# DeepPheWAS: an R package for phenotype generation and association analysis for phenome-wide association studies

**DOI:** 10.1101/2022.05.05.22274419

**Authors:** R Packer, AT Williams, W Hennah, MT Eisenberg, KA Fawcett, W Pearson, A Guyatt, A Edris, EJ Hollox, BS Rao, JR Bratty, LV Wain, F Dudbridge, MD Tobin

## Abstract

**Summary:** DeepPheWAS is an R package for phenome wide association studies that creates clinically-curated composite phenotypes, and integrates quantitative phenotypes from primary care data, longitudinal trajectories of quantitative measures, disease progression, and drug response phenotypes. Tools are provided for efficient analysis of association with any genetic input, under any genetic model, with optional sex-stratified analysis, and for developing novel phenotypes.

**Availability and Implementation:** The DeepPheWAS R package is freely available under GNU general public licence v3.0 from at https://github.com/Richard-Packer/DeepPheWAS.

**Supplementary information:** Supplementary methods and results are available at *Bioinformatics* online.

## Introduction

Phenome-wide association studies (PheWASs) can be used to better understand the pleiotropic effects of genetic variants (Tyler *et al*., 2016) and to inform drug development through target identification, target validation and use of variants that mimic drug effects to assess likely drug efficacy, safety and drug repurposing opportunities (Gill *et al*., 2019; Diogo *et al*., 2018; Khosravi *et al*., 2019). PheWASs comprise two stages – phenotype generation and statistical association tests. There have been two widely applicable methods for phenotype generation: PHESANT (Millard *et al*., 2018) and PheWAS-R (Carroll *et al*., 2014). PHESANT creates phenotypes by extracting study-specific questionnaire and measurement data alongside linked hospital records in UK Biobank. PheWAS-R combines related international classification of disease version 9 and 10 (ICD-9/ICD-10) codes into clinically relevant groups termed phecodes. Both tools provide regression analysis for per-variant PheWAS for generated phenotypes using generalised linear models in R and produce Manhattan plots. Online PheWAS resources such as Open Targets Genetics (Ghoussaini *et al*., 2021) do not perform new statistical tests. Instead, they are repositories for existing results from phenotypes generated by one of the two above tools or by individual genome wide association studies (GWAS).

These tools are useful, but have several key gaps:

1. The phenotypes generated rely on a single data field or coding ontology and do not take advantage of all available data, such as primary care data;
2. Existing approaches do not provide tools for developing new phenotypes;
3. For running per-variant PheWAS, running each regression model in R is computationally inefficient and can result in inflated type I error for low-frequency variants with a case-control imbalance (Ma *et al*., 2013);
4. Online resources such as Open Targets Genetics have limited flexibility. For example, they accept only single nucleotide polymorphisms (SNPs), and retrieve results only for genetic models tested. The user cannot specify when to use new data fields or updates to existing data fields (for example, updated health records), and the user cannot specify their preferred statistical approach and outputs, such as false discovery rate (FDR).

The platform we have developed, DeepPheWAS, addresses both phenotype generation and efficient association testing while incorporating the following developments that are not yet available in any current platform or online resource:

i. Clinically-curated composite phenotypes for selected health conditions that integrate different data types (including primary and secondary care data) to study phenotypes not well captured by current classification trees;
ii. Integration of quantitative phenotypes from primary care data, such as pathology records and clinical measures;
iii. Integration of disease progression phenotypes, longitudinal trajectories of quantitative measures and drug response measures;
iv. Clinically-curated phenotype selection for traits that are extremely highly correlated;
v. Flexible tests of additive, dominant, recessive and genotypic models;
vi. Inclusion of complex structural variants, such as multi-allelic CNVs;
vii. Ability to test genetic risk scores;
viii. Creation of phenotypes in sex-specific strata to run a sex-stratified PheWAS;
ix. Providing tools for generating novel phenotypes using a simple phenotype mapping process.

## Application of DeepPheWAS to UK Biobank

### Analysis of quantitative phenotypes

Our platform can be applied to quantitative phenotypes derived from numerous data sources, including primary care data. For example, we created a phenotype using recorded levels of blood sodium in primary care records that is not yet included in any PheWAS platform. We applied DeepPheWAS to rs7193778 (nearest genes *NFAT5* and *TERF2*), previously associated with urate levels (Köttgen *et al*., 2013). Our PheWAS shows various associations which are currently not documented in GWAS Catalog, most strongly with blood sodium levels (Supplementary Figure 1, Supplementary Table 1).

### Highly correlated traits

We applied our DeepPheWAS approach to rs2912062 (nearest genes *ANGPT2* and *AGPAT5*), shown to be associated with carotid intima-media thickness (IMT) (Strawbridge *et al*., 2020), a phenotype not currently available in any PheWAS platform. By selecting a single representative measure taken from many individual measurements DeepPheWAS can collapse highly correlated quantitative traits into single measures (in this case carotid IMT maximum and carotid IMT mean), reducing redundancy and improving power. We recapitulated known GWAS findings (Supplementary Figure 2, Supplementary Table 2).

### Flexibility with choice of genetic model

To demonstrate the flexibility of DeepPheWAS we assessed the *SERPINA1* Z allele (rs28929474(T)) under additive and recessive genetic models. Patterns of association differed between models, including opposite estimated effect directions for this variant on forced expiratory volume in 1 second (FEV_1_) and forced vital capacity (FVC), consistent with previous findings (Fawcett, Song, *et al*., 2021) (Supplementary Figures 3 and 4, Supplementary Tables 3 and 4).

### Association tests for complex structural variation

Human genomic variation includes variants which have more categories than SNPs. For example, diploid human copy number of *CCL3L1* ranges from 0 to 8 in UK Biobank participants (Fawcett, Demidov, *et al*., 2021)(Supplementary Figure 5). In such situations, association testing may be based on the measured copy number or on user-specified collapsed categories, requiring a flexible platform. We used DeepPheWAS to test association with *CCL3L1* copy number (coded 0-8) under a linear additive model); no associations reached an FDR threshold of 1% (Supplementary Figure 6, the top 5 associations are shown in Supplementary Table 5), this recapitulates findings from earlier studies (Field *et al*., 2009; Urban *et al*., 2009; Carpenter *et al*., 2011; Adewoye *et al*., 2018).

### Genetic risk scores, composite, and disease-progression phenotypes

Genetic risk scores (GRS) aggregate multiple SNPs, providing improved power for studying phenotypic associations, but cannot be specified in online PheWAS platforms. We performed a PheWAS using a 279-variant GRS for FEV_1_/FVC (Shrine N, *et al*. 2019), which showed association (FDR<1%) with 47 traits including increased risk of clinical COPD and clinical asthma with a higher score of FEV_1_/FVC reducing alleles (Supplementary Figure 7, Supplementary table 6). Furthermore, the composite phenotypes generated by the DeepPheWAS platform (for example P2020 Asthma and P2054 COPD) were consistently more strongly associated with the GRS for FEV_1_/FVC than the relevant Phecodes alone. We also show significant association with the novel disease-progression phenotypes: exacerbation of COPD and age-of-onset of COPD both of which are unavailable in existing PheWAS resources and have published GWAS results.

### Sex-stratified PheWAS

We applied DeepPheWAS to rs12777332 (nearest genes *CASP7* and *NRAP*) and rs7697189 (nearest gene *HHIP*). We replicate association for risk of cataract in women only (Supplementary Figure 8, Supplementary Table 7) for rs12777332 (Choquet *et al*., 2021), and for increased effect on FEV_1_ in men compared to women (Supplementary Figures 9 and 10, Supplementary Table 8) for rs7697189 (Fawcett, Obeidat, *et al*., 2021).

## Implementation

DeepPheWAS is an R package that can be run on high-performance computing clusters and requires R 4.1.0 and PLINK 2.0. DeepPheWAS is optimised for UK Biobank data and is expected to be interoperable with the UK Biobank Research Analysis Platform but can also be applied to other data sources see user guide.

## Availability

The DeepPheWAS R package is freely available under GNU general public licence v3.0 from at https://github.com/Richard-Packer/DeepPheWAS.

## Conclusion

Here, we present DeepPheWAS, an R package that facilitates phenome-wide association studies while addressing several limitations of existing approaches. This includes the ability to analyse a broader range of phenotypes derived from large-scale electronic healthcare records, more informative composite phenotypes, greater flexibility in the type of genetic variation that can be studied and assessing associations with genetic risk scores.

## Supporting information

User guide

## Data Availability

The raw data is available on application to UK Biobank. Scripts to produce the results highlighted here are availble from provided github address.

https://github.com/Richard-Packer/Deep-PheWAS

## Acknowledgements

This research has been conducted using UK Biobank Resource under Application Number 43027. This research used the SPECTRE High Performance Computing Facility at the University of Leicester.

## Data availability

The data underlying this article is available from the UK Biobank to all approved researchers https://www.ukbiobank.ac.uk/. Data was accessed under approved application 43027.

