## Supplementary material for "DeepPheWAS: an R package for phenotype generation and association analysis for phenome-wide association studies": User guide

**Title**: DeepPheWAS user guide

**Version**: 0.2.0

**Date**: 10/08/2022

**Author**: Richard Packer <>

**Description**: An R package for conducting large-scale phenome-wide association studies (PheWAS). For detailed information, please refer to the manuscript (doi: TBC).

**Maintainer**: Richard Packer <>

**Depends**: R(>=4.1.0), PLINK 2.0, bgenix

**Licence**: GNU general public licence v3.0

**Date/Publication**: 10/08/2022

### Introduction

DeepPheWAS is an R package designed to be used via the inbuilt R scripts. These R-scripts are accessed via the command line using a bash interface and utilise an argument parser (docopt) to process command line inputs (arguments). Docopt requires precise inputs to work and will not give any error message to highlight which input has been inputted incorrectly. It is recommended you read the description of inputs below before attempting to run the scripts.

Each script has 6 components.

1. **Overview** – a description of the overall functionality of the script.
2. **Usage** – the relationship between arguments including which arguments are required. Some scripts will have two or more usage sections which indicate different ways of using the scripts, often dependent on different inputs. The usage section follows the notation of docopt (see http://docopt.org/ for more details). In brief, arguments that are required are surrounded by () and optional arguments are surrounded by []. Where only one of several arguments should be used, these are separated by a |. For example, (arg1 | arg2) would indicate that one of arg1 or arg2 is required but not both.
3. **Arguments** – descriptions of each of the individual arguments that can be inputted into the scripts.
4. **Input** – description of the expected format of any input files including column names and any specific formatting requirements.
5. **Output** – description of the expected output, column names are not described, details can be found where the outputs are used as inputs in future scripts.
6. **Examples** – examples of the required arguments required to run the script.

#### Order to run the scripts

DeepPheWAS was designed to separate phenotype generation from association testing splitting PheWAS into smaller editable components. While it has been optimised for UK Biobank data, many of the scripts are adaptable to non-UK Biobank data sets. The inbuilt R-scripts have been split into two folders to represent the two main stages of PheWAS, phenotype generation and association testing. The R scripts are located within the DeepPheWAS package at:

/extdata/scripts/phenotype_generation/

and:

/extdata/scripts/association_testing/

The scripts have been numbered to reflect the order in which they should be run. Where scripts have the same starting number followed by a numeral they can be run in parallel.

**Stage 1: Phenotype generation**

`01_minimum_data.R`

`02_data_preparation.R`

`03a_phecode_generation.R`

`03b_creating_concepts.R`

`03c_data_field_phenotypes.R`

`03d_primary_care_quantitative_phenotypes.R`

04-formula_phenotypes.R

05_composite_phenotypes.R

**Stage 2: Association testing**

01_phenotype_preparation.R

02_extracting_snps.R**

03a_plink_association_testing.R**

03b_R_association_testing.R*

04_combine_split.R***

05_tables_graphs.R

** Only used if testing single nucleotide polymorphisms (SNPs) or other variants that can be stored in binary genetic files.

*** Only to be used the split_group option was required for 03a_plink_association_testing.R.

* Only used if testing genetic instrument that cannot be stored in binary genetic files.

### Function description

#### Phenotype generation

##### 01_minimum_data.R

**Overview**

This script combines data-field UK Biobank data—collected at the assessment centre—into a single "minimum_tab_data.gz" file that is used throughout by DeepPheWAS functions. This is achieved by subsetting the aforementioned data to columns of interest and, where the data are spread across multiple files, merging across all files provided. For UK Biobank data, the column names are referenced by data-field IDs. The desired subset of columns to be extracted is listed in the data_field_ID file found, by default, in data/fields-minimum.txt. This file can be edited to extract a different subset of columns to that provided by default. Files can be inputted in a single folder using the `data_folder` flag, or specified individually using the `data-files` flag.

**Usage**

```
01_minimum_data.R (--data_folder=<FOLDER> | --data_files=<FILES>) (--
save_loc=<FILE>) [--r_format --data_field_ID=<FILE> --
data_name_pattern=<text> --N_cores=<number> --exclusions=<FILE>]
```

**Arguments**

`save_loc` (required): Full path to the save file location for the output.

`data_folder` (required OR `data_files`): Full path of the directory that contains the data files that will be formatted and concatenated.

`data_files` (required OR `data_folder`): Comma separated full file paths of the data files that will be formatted and concatenated.

`r_format` (optional): Specific to UK Biobank data. Specify if the input for the data have been downloaded using the R option. If the data have been downloaded using the .csv or .txt options, then no input is required (default).

`data_field_ID` (optional): Full path of the file containing the field_IDs required for Deep PheWAS. Defaults to the fields-minimum.txt file located in the data/ folder.

`data_name_pattern` (optional): Character string for isolating data files if these are in a directory with other files. Defaults to using all files in the directory specified by the data_folder argument.

`N_cores` (optional): Number of cores requested if parallel computing is desired. Defaults to single core computing.

`exclusions` (optional): Full path to the file containing individuals to be excluded from the analysis. Defaults behaviour is to retain all individuals.

**Input**
Each of the data files should comprise one column of participant IDs (labelled "eid") to be used for merging, and any number of additional columns relating to data fields of interest from UK Biobank.

**Output**
This script outputs a single, tab-separated file that contains one row for each individual and as many columns as listed in the file provided by the `data_field_ID` argument.

**Examples**
1. With all optional arguments as default:

```
01_minimum_data.R \

--save_loc /path/to/save_location/file.txt \

--data_folder /path/to/data_folder/
```

2. With all arguments specified:

```
01_minimum_data.R \

--data_folder /path/to/data_folder/ \

--data_field_ID /path/to/field_ids_file.txt \

--data_name_pattern .tab.gz \

--N_cores 4 \

--save_loc /path/to/save_location/file.txt \

--exclusions /path/to/exclusions/file.txt
```

OR

```
01_minimum_data.R \

--data_files /path/to/file_1/,/path/to.file_2 \

--data_field_ID /path/to/field_ids_file.txt \

--N_cores 4 \
```

```
--save_loc /path/to/save_location/file.txt \
--exclusions /path/to/exclusions/file.txt
```

##### 02_data_preparation.R

**Overview**

This script concatenates and edits linked healthcare data from UK Biobank into a single long file with participant identifier, clinical code, date of clinical code's recording, and data source. Current data sources are:

- "MD" – when ICD-10 code is taken from mortality data.
- "cancer" – when ICD-10 code is taken from cancer registry data.
- "ICD10-1" – when an ICD-10 code is taken from hospital records as one of the primary causes of admission.
- "ICD10-2" – when an ICD-10 code is taken from hospital records as a secondary cause of admission.
- "ICD10-3" – when an ICD-10 code is taken from hospital records as an external cause of admission.
- "ICD9-1" – when an ICD-9 code is taken from hospital records as one of the primary causes of admission.
- "ICD9-2" – when an ICD-9 code is taken from hospital records as an external cause of admission.
- "ICD9-3" – when an ICD-9 code is taken from hospital records as one of the primary causes of admission.
- "OPCS" – A OPCS-4 code taken from the hospital records.
- "SR" – A self-report code for a non-cancer illness taken from data field 20002.
- "SROP" - A self-report code for an operation taken from data field 20004.
- "V2" – A Read V2 code taken from the primary care clinical data.
- "V3" – A Read V3 code taken from the primary care clinical data.

The current iteration of this script is purpose-built for UK Biobank data. As such, if this script is to be applied to non-UK Biobank data, it would first need to be formatted to mimic that of UK Biobank or to mimic the expected output of the script. The two self-reported data sources are specific to UK Biobank and if similar data were available in another data set, these would need to be mapped to the UK Biobank code used in data fields 20002 and 20004.

Primary care prescription data from UK Biobank does not lend itself to be easily combined in this way and so it is edited and saved as a separate file.

**Usage**

```
02_data_preparation.R (--save_location=<FOLDER> ) [--min_data=<FILE> --
GPC=<FILE> --GPP=<FILE> --hesin_diag=<FILE> --HESIN=<FILE> --
hesin_oper=<FILE> --death_cause=<FILE> --death=<FILE> --exclusions=<FILE>
--king_coef=<FILE>]
```

**Arguments**

`save_location` (required): Full file path for the common folder to save created files.

`min_data` (optional): Full path of the file generated by the previous step (`01_minimum_data.R`).

GPC (optional): Full path of the primary care clinical data from UK Biobank.

GPP (optional): Full path of the primary care prescription data from UK Biobank.

`hesin_diag` (optional): Full path of the hesin_diag file from UK Biobank.

`HESIN` (optional): Full path of the HESIN file from UK Biobank.

`hesin_oper` (optional): Full path of the hesin_oper file from UK Biobank.

`death_cause` (optional): Full path of the death_cause file from UK Biobank.

`death` (optional): Full path of a file containing information on date of death, where applicable.

`exclusions` (optional): Full path of the file containing individuals to be excluded from the analysis. Defaults behaviour is to retain all individuals.

`king_coef` (optional): Full file path of related data file containing King coefficient scores for related ID pairs from UK Biobank.

**Input**
This script accepts the following as input:

1. min_data: A tab-separated file containing one row of data per individual and one column for each of the phenotypes extracted from the baseline recruitment data (generated by the `01_minimum_data.R` script).
2. GPC: A tab-separated file containing the primary care clinical data that has the following columns.

**eid**: Participant identifier.
**data_provider**: Value between 1 and 4 (inclusive) that indicates the data provider—1=England (Vision), 2=Scotland, 3=England (TPP), 4=Wales.
**event_dt**: Date the clinical code was recorded.
**read_2**: Read version 2 (Read v2) code.
**read_3**: Read version 3 (Read v3 or CTV3) code.
**value1**: First recorded measurement.
**value2**: Second recorded measurement.
**value3**: Third recorded measurement.

3. GPP: A tab-separated file containing the primary care prescription data that has the following columns.

**eid**: Participant identifier.
**data_provider**: Value between 1 and 4 (inclusive) that indicates the data provider—1=England (Vision), 2=Scotland, 3=England (TPP), 4=Wales.
**issue_date**: Date the prescription was issued.
**read_2**: Read versions (Read v2) code.
**bnf_code**: British National Formulary (BNF) code.
**dmd_code**: Dictionary of medicines and devices (dmd) code.
**drug_name**: Description of the prescription issued.
**quantity**: Description of the quantity issued.

4. hesin_diag: A tab-separated file containing in-patient hospital records (secondary care) that has the following columns.

**eid**: Participant identifier.
**ins_index**: Instance index that denotes a unique record when combined with the participant identifier.
**arr_index**: A sequential index, starting at 0, which labels each separate diagnosis.

**level**: Value between 1 and 3 (inclusive)—1=main diagnosis, 2=secondary diagnosis, 3=external cause.
**diag_icd9**: ICD-9 code.
**diag_icd9_nb**: ICD-9 code addendum.
**diag_icd10**: ICD-10 code.
**diag_icd10_nb**: ICD-10 code addendum.

5. HESIN: A tab-separated file containing in-patient hospital administrative data that is described in more detail by downloading the HESDataDic.xlsx file available at https://biobank.ctsu.ox.ac.uk/crystal/refer.cgi?id=141140.
6. hesin_oper: A tab-separated file containing operation data that has the following columns.

**eid**: Participant identifier.
**ins_index**: Instance index that denotes a unique record when combined with the participant identifier.
**arr_index**: A sequential index, starting at 0, which labels each separate procedure.
**level**: Value between 1 and 2 (inclusive)—1=main operation, 2=secondary operation.
**opdate**: Date of operation.
**oper3**: OPCS-3 code.
**oper3_nb**: OPCS-3 code note.
**oper4**: OPCS-4 code.
**oper4_nb**: OPCD-4 code note.
**posopdur**: Duration of post-operative stay in hospital.
**preopdur**: Duration of pre-operative stay in hospital.

7. death_cause: A tab-separated file containing cause of death information that has the following columns.

**eid**: Participant identifier.
**ins_index**: Instance index that denotes a unique record when combined with the participant identifier.
**arr_index**: A sequential index, starting at 0, which labels each separate cause of death.
**level**: Value between 1 and 2 (inclusive)—1=primary cause of death, 2=contributory cause of death.
**cause_icd10**: ICD-10 code.

8. death: A tab-separated file containing date of death that has the following columns.

**eid**: Participant identifier.
**ins_index**: Instance index that denotes a unique record when combined with the participant identifier.
**dsource**: Provider of the death information whose coding is described at https://biobank.ctsu.ox.ac.uk/crystal/coding.cgi?id=1970.
**source**: Death information source whose coding is described at https://biobank.ctsu.ox.ac.uk/crystal/coding.cgi?id=261.
**date_of_death**: Date of death.

9. exclusions: A file containing a single column of participant identifiers to be excluded, no header.
10. king_coef: A white space-separated file that has the following columns.

**ID1**: Identifier for first participant in pair.
**ID2**: Identifier for second participant in pair.
**Kinship**: Kinship coefficient estimate.

**Output**

This script outputs the following files:

1. combined_sex: A tab-separated file that contains one column of participant IDs and another column that describes participant sex--obtained from Data-Field 31 and Data-Field 22001. Data-Field 31 is only used where Data-Field 22001 is missing.
2. control_exclusions: A file that contains a column of participants IDs corresponding to participants who have been lost to follow-up (Data-Field 190).
3. related_callrate: A tab-separated file that describes all pairs of individuals with a kinship coefficient from the KING software exceeding 0.0884 (more distantly related than second-degree relatives) combined with call rate data from genetic quality control.
4. GP_C_edit.txt.gz: A lightly edited version of the UK Biobank primary care clinical data with individuals listed in the `exclusions` argument removed and the date a record was made reformatted.
5. GP_C_ID.txt.gz: A file that lists participants IDs according to those present in the UK Biobank primary care clinical data.
6. GP_P_edit.txt.gz: A lightly edited version of the UK Biobank primary care prescription data with individuals listed in the `exclusions` argument removed and the date a prescription was given reformatted.
7. health_records.txt.gz: A tab-separated file that contains hospital in-patient data (Hospital Episode Statistics), operations and procedures data and mortality data, one row of data per data point and individual.
8. control_populations: A tab-separated file that contains IDs of all participants that have records of sex recorded and participants with a GP record. Used to define control populations for the composite phenotypes.

**Examples**

1. With no arguments specified (this script produces no output if run this way):

```
02_data_preparation.R \

--save_location /path/to/save_folder/
```

2. With all arguments specified (generates all phenotypes specified in data/PheWAS_manifest.csv):

```
02_data_preparation.R\

--min_data /data/minimum_tab_data.gz \

--GPC /path/to/primarycare_clinical/file.txt \

--GPP /path/to/primarycare_prescription/file.txt \

--hesin_diag /path/to/hesin_diag/file.txt \

--HESIN /path/to/HESIN/file.txt \

--hesin_oper /path/to/hesin_oper/file.txt \

--death_cause /path/to/cause_of_death/file.txt \

--death /path/to/date_of_death/file.txt \

--exclusions /path/to/exclusions/file.txt \

--king_coef /path/to/king_relatedness_info/file.txt \
```

```
--save_location /path/to/save_folder/
```

#### 03a_phecode_generation.R

**Overview**

This script maps healthcare data from the ICD-9 and ICD-10 coding frameworks to Phecodes (https://phewascatalog.org/phecodes) and generates output that describes the resulting phenotypes in all individuals. In addition, output can be generated that describes individuals who should be excluded when selecting controls (binary phenotypes). For example, if one were interested in defining an asthma phenotype but wanted any controls to not have chronic obstructive pulmonary disease codes, that can be done using this script.

**Usage**

```
03a_phecode_generation.R (--health_data=<FILE> --sex_info=<FILE>) (--
phecode_save_file=<FILE> | --no_phecodes) (--range_ID_save_file=<FILE> | -
-no_range_ID) [--control_exclusions=<FILE> --N_cores=<number>
--ICD10=<text_comma> --ICD9=<text_comma>]
```

**Arguments**

`health_data` (required): Full path of the file produced by the previous step (`02_data_preparation.R`).

`sex_info` (required): Full path of the "combined_sex" file produced by the previous step.

`phecode_save_file` (required if not using `no_phecodes`): Full path of the save file for the phecode phenotypes R data object.

`range_ID_save_file` (required if not using `no_range_ID`): Full path of the folder used to store the control exclusions R data object.

`control_exclusions` (optional): Full file path of the optional control exclusions file.

`N_cores` (optional): Number of cores requested if parallel computing is desired. Defaults to single core computing.

`no_range_ID` (optional): Choose whether to save lists of participant identifiers to be excluded from control definitions (FALSE) or not (TRUE). Defaults to FALSE.

`no_phecodes` (optional): Choose whether to save the Phecode-generated phenotype data (FALSE) or not (TRUE). Defaults to FALSE.

`ICD10` (optional): Comma-separated string representing the column names used in `health_data` for ICD-10 values. If there are no ICD-10 values, use NA or any text not used as a source in health data. Defaults to "ICD10".

`ICD9` (optional): Comma-separated string representing the column names used in `health_data` for ICD-9 values. If there are no ICD-9 values, use NA or any text not used as a source in health data. Defaults to "ICD9".

**Input**

This script accepts the following files as input:

health_records.txt.gz (default name): A tab-separated file that contains hospital in-patient data (Hospital Episode Statistics), operations and procedures data and mortality data, one row of data per data point and individual. This is generated by the `02_data_preparation.R` script.

1. combined_sex (default name): A tab-separated file that contains one column of participant IDs and another column that describes participant sex. This is used for sex-stratified phenotypes.
2. control_exclusions (default name): A file with a single column that describes the IDs of participants who will not be used as controls for binary phenotypes.

**Output**
This script outputs the following files:

1. phecodes.RDS (default name): R data object containing a list of dataframes. Each dataframe describes the Phecode-mapped phenotypes in all individuals and has the following three columns.

**eid**: Participant identifier.
**phecode**: Frequency of the corresponding Phecode in the participant's healthcare data.
**earliest_date**: Date of first instance of the corresponding Phecode for the participant.

2. range_ID.RDS (default name): R data object containing a list of dataframes. Each dataframe gives participant identifiers for those who should be excluded when selecting controls (binary phenotypes).

**Examples**
1. With only required arguments specified:

```
03a_phecode_generation.R \

--health_data /path/to/health_data/health_records.txt.gz \

--sex_info /path/to/sex_data/combined_sex

--phecode_save_file /path/to/phecode_save_location

--range_ID_save_file /path/to/phecode_save_location
```

##### 03b_data_field_phenotypes.R

**Overview**
This script creates Data-Field phenotypes from UK Biobank Data-Field IDs and primary care data. The script uses the PheWAS_manifest.csv file (provided with the Platform, located in the data/ folder) as a guide to creating phenotypes and will only create phenotypes where the required Data-Field ID data is available. If data from outside UK Biobank are being used, UK Biobank Data-Field IDs would need to be mapped to their equivalent in the alternative data source.

**Usage**
```
03b_data_field_phenotypes.R (--min_data=<FILE> --
phenotype_save_file=<FILE>) [--N_cores=<number> --
PheWAS_manifest_overide=<FILE> --append_file=<FILE>]
```

**Arguments**
`min_data` (<u>required</u>): Full path of the tab-separated file generated by the `01_minimum_data.R` script.

`phenotype_save_file` (<u>required</u>): Full path for the save file for the generated data field phenotypes RDS to be used for phenotype creation.

`N_cores` (optional): Number of cores requested if parallel computing is desired. Defaults to single core computing.

PheWAS_manifest_overide (optional): Full path of an alternative to the in-built file.

append_file (optional): Full path of an existing R object containing Data-Field phenotype information to which additional phenotypes derived by this script will be appended.

**Input**
This script accepts the following files as input:

1. minimum_tab_data.gz (default name): The single, tab-separated file that is output by the 01_minimum_data.R script. The file has one row per individual and one column per phenotype of interest (derived from the UK Biobank Data-Field data).
2. alternative_manifest.csv: An alternative PheWAS manifest to that provided with the Platform.

**Output**

This script outputs the following files:

1. data_field_phenotypes.RDS (any name inputted into phenotype_save_file): R data object containing a list of dataframes. Each dataframe describes Data-Field phenotypes derived from UK Biobank Data-Field IDs and primary care data.

**Examples**
1. With only required arguments specified:

```
03b_data_field_phenotypes.R \

--min_data /data/minimum_tab_data.gz

--phenotype_save_file /data/phenotypes/data_field_phenotypes.RDS
```

2. With all arguments specified:

```
03b_data_field_phenotypes.R \

--min_data /data/minimum_tab_data.gz \

--N_cores 12 \

--PheWAS_manifest_overide /path/to/alternative_manifest/file.txt \

--phenotype_save_file /data/phenotypes/data_field_phenotypes.RDS \

--append_file /path/to/existing_data/file.RDS
```

#### 03c_creating_concepts.R

**Overview**
This script uses code lists provided to define concepts. Concepts can then be combined to form cases and controls for "composite-phenotypes". A concept is a single homogenous group of codes that all describe a single disease, symptom, or related group of medicines.

The main output that is used for forming phenotypes is saved as an R object and is a record of the ID, number of codes and the date of the earliest code in a record. The other files are used for creating summary documents of the concepts.

UK Biobank prescription data is unusual in that it does not contain consistent codes (namely BNF codes) for most of the data. As such, much of the searching is done using key search terms. A small subsection requires using Read version 2 drug codes. Because of this major difference with the other types of data, prescription concepts have their own function and would need to be used with caution in a non-UK Biobank setting.

Users can choose to define only concepts that use clinical data or prescription data or both. At least one must be inputted, however.

**Usage**

```
03c_creating_concepts.R (--GPP=<FILE> concept_save_file=<FILE> --
all_dates_save_file=<FILE>) [--health_data=<FILE> --
PheWAS_manifest_overide=<FILE> --code_list_folder=<FOLDER>]
```

**OR**

```
03c_creating_concepts.R (--health_data=<FILE> --concept_save_file=<FILE> -
-all_dates_save_file=<FILE>) [--GPP=<FILE> --
PheWAS_manifest_overide=<FILE> --code_list_folder=<FOLDER>]
```

**Arguments**

GPP (required if `health_data` argument is not specified): Full path of the primary care prescription data from UK Biobank. Specifying this argument will generate prescription concepts.

`health_data` (required if GPP argument is not specified): Full path of the heath_data file for UK Biobank. Specifying this argument will generate clinical concepts.

`concept_save_file` (required): Full path of the save file for the generated concepts RDS to be used for phenotype creation.

`all_dates_save_file` (required): Full file path for the save file for the generated all_dates.RDS used for per-event combinations of concepts.

`PheWAS_manifest_overide` (optional): Full path of an alternative to the inbuilt PheWAS_manifest file.

`code_list_folder` (optional): Full path of the folder containing code lists. Only use if not using default stored in R package.

**Input**

This script accepts the following files as input

1. health_records.txt.gz (default name): A tab-separated file that contains hospital in-patient data (Hospital Episode Statistics), operations and procedures data and mortality data, one row of data per data point and individual. This is generated by the `02_data_preparation.R` script.
2. GP_P_edit.txt.gz (default name): A lightly edited version of the UK Biobank primary care prescription data. This file is generated by the `02_data_preparation.R` script.
3. alternative_manifest.csv: An alternative PheWAS manifest to that provided with the Platform.

**Output**

This script outputs the following files:

1. concepts.RDS (any name inputted into `concept_save_file`): R data object containing a list of dataframes. Each dataframe describes the concept-based phenotypes in all individuals and has the following three columns.

**eid**: Participant identifier.

**concept_code**: Frequency of the corresponding concept code in the participant's healthcare data.
**earliest_date**: Date of first instance of the corresponding concept code for the participant.

2. all_dates.RDS (default name): R data object containing a list of dataframes. Each dataframe gives the date the corresponding concept was recorded in an individual's primary care clinical or prescription data.

**Examples**

1. Requesting prescription-based concepts only:

```
03c_creating_concepts.R \
--GPP /path/to/primarycare_prescription/file.txt
--concept_save_file /path/to/concept_save_file \
--all_dates_save_file /path/to/all_dates_save_file
```

2. Requesting clinical-based concepts only:

```
03c_creating_concepts.R \
--health_data /path/to/primarycare_clinical/file.txt
--concept_save_file /path/to/concept_save_file \
--all_dates_save_file /path/to/all_dates_save_file
```

3. Requesting both prescription-based and clinical-based concepts:

```
03c_creating_concepts.R \
--GPP /path/to/primarycare_prescription/file.txt \
--health_data /path/to/primarycare_clinical/file.txt \
--concept_save_file /path/to/concept_save_file \
--all_dates_save_file /path/to/all_dates_save_file
```

#### 03d_primary_care_quantitative_phenotypes.R

**Overview**

This script creates quantitative phenotypes from UK Biobank primary care data. The script uses the PheWAS_manifest.csv file (provided with package) as a guide to creating phenotypes.

**Usage**

```
03_dprimary_care_quantitiative_phenotypes.R (--GPC=<FILE> --DOB=<FILE>
phenotype_save_file=<FILE>) [--N_cores=<number> --
PheWAS_manifest_overide=<FILE> --]
```

**Arguments**

GPC (required): Full path of the primary care clinical data file from UK Biobank.

DOB (required): Full path of a file describing participant date of birth. This does not need to be the exact date. By default for UK Biobank data, the script creates the date of birth from month and year of birth (data-fields 52 and 34).

`phenotype_save_file` (required): Full path of the save file for the generated primary care quantitative phenotypes RDS to be used for phenotype creation.

`N_cores` (optional): Number of cores requested if parallel computing is desired. Defaults to single core computing.

`PheWAS_manifest_overide` (optional): Full file path of the alternative PheWAS_manifest file.

**Input**

This script accepts the following files as input:

1. GP_C_edit.txt.gz (default name): A lightly edited version of the UK Biobank primary care clinical data. This file is generated by the `02_data_preparation.R` script.
2. DOB: A comma-separated file that has column names "eid" and "DOB", the former listing participant identifiers and the latter their date of birth.
3. alternative_manifest.csv: An alternative PheWAS manifest to that provided with the Platform.

**Output**

This script outputs the following files:

1. PQP.RDS (any name from argument `phenotype_save_file`): R data object containing a list of dataframes. Each dataframe describes quantitative traits derived from primary care data.

**Examples**

1. With only required arguments specified:

```
03d_primary_care_quantitiative_phenotypes.R \

--GPC /path/to/primarycare_clinical/file.txt \

--DOB /path/to/date_of_birth/file.txt \

--phenotype_save_file /data/phenotypes/quantitative_phenotypes.RDS
```

2. With all arguments specified:

```
03d_primary_care_quantitiative_phenotypes.R \

--GPC /path/to/primarycare_clinical/file.txt \

--DOB /path/to/date_of_birth/file.txt \

--phenotype_save_file /data/phenotypes/quantitative_phenotypes.RDS \

--N_cores 12 \

--PheWAS_manifest_overide /path/to/alternative_manifest/file.txt
```

##### 04_formula_phenotypes.R

**Overview**

This script creates formula phenotypes from the phenotype data generated by earlier scripts. The script uses the PheWAS_manifest.csv file (provided with the package) as a guide to creating phenotypes.

**Usage**

```
04_formula_phenotypes.R (--min_data=<FILE> --phenotype_save_file=<FILE>)
[--data_field_phenotypes=<FILE> --sex_info=<FILE> --
PheWAS_manifest_overide=<FILE>]
```

**Arguments**
`min_data` (required): Full path of the file generated by the previous step (`01_minimum_data.R`).

`phenotype_save_file` (required): Full path of the save file for the generated formula phenotype RDS to be used for phenotype creation.

`data_field_phenotypes` (optional): Full path of the data_field.RDS file produced by `03b_data_field_phenotypes.R`, currently only used for eGFR phenotype.

`sex_info` (optional): Full path of the combined_sex file (generated by the 02_data_preparation.R script).

`PheWAS_manifest_overide` (optional): Full file path of the alternative PheWAS_manifest file.

**Input**

This script accepts the following files as input:

1. minimum_tab_data.gz (default name): The single, tab-separated file that is output by the `01_minimum_data.R` script. The file has one row per individual and one column per phenotype of interest (derived from the UK Biobank Data-Field data).
2. data_field.RDS: The R data object generated by the `03b_data_field_phenotypes.R` script.
3. combined_sex (default name): A tab-separated file that contains one column of participant IDs and another column that describes participant sex. This is used for sex-stratified phenotypes.
4. alternative_manifest.csv: An alternative PheWAS manifest to that provided with the Platform.

**Output**
This script outputs the following files:

1. formula_phenotypes.RDS (any name inputted into `phenotype_save_file`): R data object containing a list of dataframes. Each dataframe describes formula phenotypes derived.

**Examples**
1. With only required arguments specified:

```
04_formula_phenotypes.R \

--min_data /data/minimum_tab_data.gz \

--phenotype_save_file /data/phenotypes/formula_phenotypes.RDS
```

2. With all arguments specified:

```
04_formula_phenotypes.R \

--min_data /data/minimum_tab_data.gz \

--phenotype_save_file /data/phenotypes/formula_phenotypes.RDS \

--data_field_phenotypes /data/phenotypes/data_field_phenotypes.RDS \

--sex_info /path/to/sex_data/combined_sex \

--PheWAS_manifest_overide /path/to/alternative_manifest/file.txt
```

#### 05_composite_phenotypes.R

**Overview**

This script creates composite phenotypes and composite concepts using a central map file. Also creates population_control ID lists that are used when creating the composite phenotypes. The phenotype creating script iterates over itself (default five times) as some composite phenotypes use other composite phenotypes in their definition.

**Usage**

```
05_composite_phenotypes.R (--phenotype_folder=<FOLDER> | --
phenotype_files) (--phenotype_save_file=<FILE>)  [--
composite_phenotype_map_overide =<FILE> --control_populations=<FILE> --
N_iterations=<number> --update_list]
```

**Arguments**

`phenotype_save_file` (required): Full path for the save file for the generated composite phenotypes RDS.

`phenotype_folder` (required if not using phenotype_files): Full path of the folder containing the phenotype data created in previous steps.

`phenotype_files` (required if not using phenotype_folder): Comma separated full file paths of phenotype data created in previous steps.

`composite_phenotype_map_overide` (optional): Full path of the "composite_phenotyope_map" file. Provided with the package.

`control_populations` (optional): Full path of the "control_populations" file containing columns of IDs, column names are used to create lists of IDs. Saved as a list called control_populations.RDS in the folder inputted in the phenotype_folder flag. The lists are used to define some of the composite phenotype control populations as directed by the composite_phenotype_map file. The unedited version of the composite_phenotype_map file uses two populations, all_pop and primary_care_pop, which represent all IDs in the sample and all IDs with available primary care data. To create composite phenotypes, the names of these list must match the composite_phenotype_map file.

The `02_data_preparation.R` script creates a "control_populations" file that is used by default.

`N_iterations` (optional): Number of iterations the script is to run through. Defaults to five.

`update_list` (optional): Option to run this script as an update to an existing composite_phenotype list object. If specified, this script will use `phenotype_save_file` to load the existing list and then save over that file upon completion.

**Input**

This script accepts the following files as input:

1. composite_phenotype_map.csv (default name): An alternative composite phenotypes map to that provided with the Platform. The formatting of this alternative manifest will be detailed in a later version of this user guide.
2. control_populations (default name): A tab-separated file that describes individuals to be used as controls for binary phenotypes. This file is automatically generated by the `02_data_preparation.R` script.

**Output**

This script outputs the following files:

1. composite_phenotypes.RDS (any name inputted into `phenotype_save_file`): R data object containing a list of dataframes. Each dataframe describes the composite phenotypes in all individuals and has the following three columns.

**eid**: Participant identifier.
**any_code**: Frequency of the codes comprising the composite phenotype in the participant's healthcare data.
**earliest_date**: Date of first instance of the corresponding concept code for the participant.

**Examples**
1. With minimal arguments specified:

```
05_composite_phenotypes.R \

--phenotype_folder /path/to/data/phenotypes/ \

--phenotype_save_file /path/to/save_location/composite_phenotypes.RDS
```

OR

```
--phenotype_files /path/to/data/phenotypes/file_1.RDS,
/path/to/data/phenotypes/file_2.RDS \

--phenotype_save_file /path/to/save_location/composite_phenotypes.RDS
```

2. With all arguments specified:

```
05_composite_phenotypes.R \

--phenotype_folder /path/to/data/phenotypes/ \

--phenotype_save_file /path/to/save_location/composite_phenotypes.RDS \

--control_populations /path/to/data/control_populations \

--N_iterations 6 \

--composite_phenotype_map_overide
/path/to/data/composite_phenotype_map.csv
```

#### Association testing scripts

##### 01_phenotype_preparation.R

**Overview**
This script takes the phenotypes generated by previous steps and filters them to those exceeding a defined number of cases and, optionally, relatedness. This filtering can be done on the whole sample or within defined groups of individuals, such as by ancestry or sex. Secondly the script reformats and then converts the phenotype data into a table prior to association testing.

**Usage**
```
phenotype_preparation.R (--phenotype_folder=<FOLDER> | --
phenotype_files=<FILES>) (--phenotype_filtered_save_name=<FILE>)  [(--
relate_remove --kinship_file=<file>)] [(--sex_split_phenotypes=<FILE> --
sex_info=<file>)] [(--age_of_onset_phenotypes=<FILE> --DOB_file=<FILE>)]
[--groupings=<file> --quantitative_Case_N=<number> --
binary_Case_N=<number> --male=<number> --female=<number> --
```

PheWAS_manifest_overide=<FILE> --IVNT --save_RDS=<FILE> --stats_save=<FILE>]

**Arguments**
phenotype_filtered_save_name (required): Full file path for save location of filtered phenotypes. Saves and dataframe per-group (if provided) that is the input to the association testing. Name of final save is /path/to/file/(group)_savename.

phenotype_folder (required if not using phenotype_files): Full path of the folder containing the phenotype data created in previous steps.

phenotype_files (required if not using phenotype_folder): Comma separated full file paths of phenotype data created in previous steps.

sex_split_phenotypes (optional): Full file path to file containing single column labelled PheWAS_ID, containing PheWAS_IDs to make sex specific phenotypes, will derive and add a male and female version of each phenotype. All sex specific phenotypes be labelled (PheWAS_ID)_male or (PheWAS_ID)_female.

sex_info (optional): Full path of the combined_sex file (generated by the 02_data_preparation.R script).

male (optional): Value corresponding to males in the combined_sex file. Default value is 1.

female (optional): Value corresponding to females in the combined_sex file. Default value is 0.

age_of_onset_phenotypes (optional): Full file path to file containing three columns PheWAS_ID,lower_limit,upper_limit,transformation. PheWAS_ID are the phenotypes to create the age_of_onset phenotypes for, lower_limit is the lower age boundary to filter read as <=, upper_limit is the upper age boundary acceptable read as >=. Transformation is the type of transformation (if any) that should be applied to the phenotype. Current accepted transformations are IVNT for inverse normal transformation. All age of onset phenotypes to be labelled (PheWAS_ID)_age_of_onset.

DOB_file (optional): Full file path to file containing DOB information.

save_RDS (optional): Full file path to save location to save and intermediate RDS file that has been filtererd by case_N and relatedness (if chosen) but not converted to dataframe for analysis.

groupings (optional): Full path of the file containing group information used for stratifying the sample. If this argument is specified, the phenotype filtering will be performed within the groups. Otherwise, the filtering will be performed in all individuals.

quantitative_Case_N (optional): The minimum number of individuals required for quantitative phenotypes. Default value is 100 individuals.

binary_Case_N (optional): The minimum number of cases required for binary phenotypes. Default value is 100 cases.

relate_remove (optional): Specify whether related individuals are additionally excluded (TRUE) or not (FALSE). Default is FALSE.

kinship_file (optional): Full path to the file containing kinship coefficient estimates.

PheWAS_manifest_overide (optional): Full file path of the alternative PheWAS_manifest file.

`IVNT` (optional): Specify whether quantitative phenotypes, defined in the PheWAS manifest, should undergo a rank-based inverse normal transformation (TRUE) or not (FALSE). Default is FALSE.

`stats_save` (optional): Full file path to save file for stats, saves summary stats for phenotypes for number of cases and controls in each grouping variable. File name will be appended with _N_filtered.csv or _relate_remove.csv, depending on if relate remove was selected.

**Input**
This script accepts the following files as input:

1. groupings: A tab-separated file that contains one column of participants IDs and another column for group assignment. The column headers should be named "eid" and "group", respectively.
2. related_callrate (default name): A tab-separated file that contains the following six columns.

**ID1**: Identifier for the first participant in the related pairing.
**ID2**: Identifier for the second participant in the related pairing.
**Kinship**: Kinship coefficient estimate for the related pairing.
**missingness_ID1**: SNP missingness (100 – SNP call rate)% for first individual in the related pairing.
**missingness_ID2**: SNP missingness (100 – SNP call rate)% for second individual in the related pairing.
**lower_missing**: Indicator of which individual has the lower SNP missingness.

3. combined_sex (default name): A tab-separated file that contains one column of participant IDs and another column that describes participant sex. This is used for sex-stratified phenotypes.
4. alternative_manifest.csv: An alternative PheWAS manifest to that provided with the Platform.

**Output**
This script outputs the following files:

1. all_pheno_epi_case_N_filtered (default name).
2. relate_remove_pheno_N_filtered (default name).

**Examples**
1. With minimal arguments specified:

```
01_phenotype_preparation.R \

--phenotype_folder /path/to/data/phenotypes/ \

--phenotype_filtered_save_name /path/to/save_name
```

2. With all arguments specified:

```
01_phenotype_preparation.R \

--phenotype_folder /path/to/data/phenotypes/ \

--phenotype_filtered_save_name /path/to/save_name \

--groupings /path/to/groupings \

--quantitative_Case_N 100 \

--binary_Case_N 50 \

--no_case_N_save FALSE \

--relate_remove TRUE \
```

```
--kinship_file /path/to/data/related_callrate \
--age_of_onset_phenotypes /path/to/age_of_onset_phenotypes \
--DOB_file /path/to/DOB_file \
--sex_split_phenotypes /path/to/sex_split_phenotypes
--sex_info /path/to/data/combined_sex \
--male 1 \
--female 0 \
--PheWAS_manifest_overide /path/to/alternative_manifest/file.txt \
--IVNT \
--stats_save /path/to/stats_save \
--save_RDS /path/to/save_RDS
```

#### 02_extracting_snps.R

**Overview**

This script uses bgenix or PLINK 2 to extract SNPs from genetic data, in .bgen, .bed or .pgen formats, using SNP identifiers and chromosome number. As such, bgenix and/or PLINK should be installed.

First, the genetic_file_guide_template.csv file must be edited to provide the file location of the genetic files and corresponding .sample/.fam/.psam file for each of the 22 autosomal and 2 sex chromosomes. If genetic data for a particular chromosome or chromosome(s) are missing, the corresponding rows in the data/genetic_file_guide_template.csv file should be deleted. If the genetic data are not stored per chromosome, then different values can be used in the chromosome column. The genetic_file_guide_template.csv is found within the R package in the extdata folder compressed as a gzip file. A template for SNP_list.csv is also saved in extdata.

**Usage**

```
02_extracting_snps.R (--genetic_file_guide=<FILE> --SNP_list=<FILE> --
analysis_folder=<FOLDER>) (--plink_input | --bgen_input) [--
plink_exe=<text> --bgenix_exe=<text> --plink_type=<text> --ref_bgen=<text>
--variant_save_name=<name> --no_delete_temp]
```

**Arguments**

`genetic_file_guide` (required): Full path of the completed genetic_file_guide_template.csv (default name) file (described in the Input section below).

`SNP_list` (required): Full path of the file describing which SNPs are to be extracted from the genetic data files ahead of association testing. The required formatting for this file is described in the Input section below.

`analysis_folder` (required): Full path of the folder that will contain the data for the SNPs given by `SNP_list`. A temporary folder, named "temp_plink" by default, will be created within the folder specified by this argument as a place to hold temporary files needed for the SNP extraction process. Unless otherwise requested by specifying the `no_delete_temp` argument, the temporary folder and its contents will be deleted following successful SNP extraction.

**One of the two following arguments must be specified.**

`plink_input` (<u>required if `bgen_input` is unused</u>): Specify that the genetic data files are in PLINK format (.bed or .pgen).

`bgen_input` (<u>required if `plink_input` is unused</u>): Specify that the genetic data files are in .bgen format.

`plink_exe` (optional): Full path to the PLINK2 executable.

`bgenix_exe` (optional): Full path to the bgenix executable.

`plink_type` (optional): Specify whether the PLINK-formatted genetic data are in .bed or .pgen format. Default is .bed.

`ref_bgen` (optional): One of the following three values that specifies which allele is to be used as the reference: ref-first (first allele is the reference, default), ref-last (last allele is the reference), red-unknown (last allele is provisionally treated as the reference).

`variant_save_name` (optional): Name of the output genetic data files. Defaults to variants_for_association.

`no_delete_temp` (optional): Specify whether the temporary folder containing intermediate files should be retained (TRUE) or deleted (FALSE). Default is TRUE.

**Input**
This script accepts the following files as input:

1. genetic_file_guide_template.csv (default name): A comma-separated file that contains one row of data per chromosome and the following three columns.

**chromosome**: Chromosome identifier, typically a number from 1 to 22 or the letters X and Y.
**genetic_file_location**: Full path of the genetic data file for the corresponding chromosome.
**psam_fam_sample_file_location**: Full path of the .sample, .fam or .pgen file, as necessary.

2. SNP_list: A comma-separated file that contains one row per SNP and the following five columns.

**chromosome:** Chromosome identifier, typically a number from 1 to 22 or the letters X and Y. This should be in the same format as the genetic_file_guide_template.csv file. For example, not expressed as "01" or "chr1" if this is given as "1" in the above file.
**rsid:** SNP identifier.
**group_name:** Option to assign a common group name to SNPs that are related, such as being in the same credible set.
**coded_allele:** Coded allele for the corresponding SNP.
**non_coded_allele:** Non-coded allele for the corresponding SNP.

**Output**
This script outputs the following files:

1. variants_for_association.pgen (default name): PLINK 2.0 binary file containing the genetic data for the variants to be studied.
2. variants_for_association.psam (default name): PLINK 2.0 row indices (individuals) corresponding to the .pgen file.
3. variants_for_association.pvar (default name): PLINK 2.0 column indices (SNPs) corresponding to the .pgen file.

**Examples**

1. With only required arguments specified:

```
02_extracting_snps.R \
--genetic_file_guide /path/to/genetic_file_guide_template.csv \
--SNP_list /path/to/SNP_list \
--analysis_folder /path/to/output_folder/ \
--plink_input
```

2. With all arguments specified:

```
02_extracting_snps.R \
--genetic_file_guide /path/to/genetic_file_guide_template.csv \
--SNP_list /path/to/SNP_list \
--analysis_folder /path/to/output_folder/ \
--plink_input \
--plink_exe /path/to/plink_executable \
--plink_type bed \
--variant_save_name variants_for_association \
--no_delete_temp
```

#### 03a_PLINK_association_testing.R

**Overview**

Runs association analysis in using PLINK 2. Takes the variants extracted using `02_extracting_snps.R` and performs regression analysis on the availble phenotypes per inputted group.

Requires location of a folder where analysis will be hosted, a comma separated list of phenotype files derived from `01_phenotype_preparation.R`, a covariate file edited for use in plink (see user guide), and the location of the variants for association prepared with `02_extracting_snps.R`. By default, it will run association tests on all included phenotypes in the phenotype files used and use those phenotype files to assign group names, all analysis is then performed per group. Phenotypes can be specified using one of two arguments phenotype_inclusion_file or phenotype_exclusion_file, group names can be specified using the group_name_overide argument. Results are saved as an R object with option to save tables per-group of the combined raw plink results. With a large number of variants the regression analysis can take a long time, to make for more efficient analysis the phenotypes for any one or more of the analysed groups can be split using the split_group input. When a group is split the phenotype files are split into smaller chunks, the analysis can then be performed in a cluster enviroment.To perfomrm the analysis this same script can be used again but with the split_analysis flag used to amend how results are saved. Results are then combined with `04_combine_split_analysis.R`. If splitting analysis tables and figures should not be compiled until `04_combine_split_analysis.R` is complete.

**Usage**

```
03a_PLINK_association_testing.R  (--analysis_folder=<FOLDER> --
covariate=<file> --phenotype_files=<FILE> --
variants_for_association=<FILE> --analysis_name=<name>) [--
group_name_overide=<text> --split_analysis --
PheWAS_manifest_overide=<FILE> --plink_exe=<command> --save_plink_tables -
-split_group=<name> --N_quant_split=<number> --N_binary_split=<number> --
model=<text> --check_existing_results] [--phenotype_inclusion_file=<FILE>
| --phenotype_exclusion_file=<FILE>]
```

**Arguments**

`analysis_folder` (required): Full path of the directory in which the association results will be saved.

`covariate` (required): Full path of a file containing covariate data to be used in model adjustment.

`phenotype_files` (required): Comma-separated list containing the full paths of the phenotype data.

`variants_for_association` (required): Full path of the genetic data for the SNPs of interest (produced by the 02_`extracting_snps.R` script).

`analysis_name` (required): Name for the analysis to be used in the naming of the output files.

`group_name_overide` (optional): Comma-separated list containing alternative group names. By default, group names are extracted from the suffix of the file names provided in the `phenotype_files` argument. For example, if a file named /home/phenotypes/EUR_phenotypes.csv were provided, the corresponding group name would be "EUR". This argument allows for a different group name to be specified. The order of names provided should match the files specified in the `phenotype_files` argument.

`split_analysis` (optional): Specify whether the input being analysed is from the `split_group` argument.

`PheWAS_manifest_overide` (optional): Full file path of the alternative PheWAS_manifest file.

`plink_exe` (optional): Full path to the PLINK 2.0 executable.

`save_plink_tables` (optional): Specify whether results of the association analysis should be saved as a table. These tables are saved in `analysis_folder`/association_results/group/group_plink_results_raw/

`split_group` (optional): Comma separated groups that require splitting for more efficient analysis. Does not run the analysis per group but splits and saves the phenotypes into smaller files which can then be analysed using cluster based computing. The number of phenotypes in each split file is dependent on the type (binary or quantitative) and is set using N_quant_split and N_binary_split. Split files are saved in analysis_folder/group_split with group being the group name. A file is created in analysis_folder/group_split saved as group_split_guide with group once again being the group name inputted. This file lists the file names of the splits, which can be used to guide distributed computing analysis. Combine the results using `04_combine_split_analysis.R`.

`N_quant_split` (optional): The number of quantitative phenotypes in each batch. Default value is 200.

`N_binary_split` (optional): The number of binary phenotypes in each batch. Default value is 80.

`model` (optional): Genetic model to use for the analysis. This can be one of genotypic, hethom, dominant, recessive, hetonly.

`check_existing_results` (optional): Specify whether part of the analysis should be re-run while checking for existing results in the `plink_results` folder. Used primarily to recover a run that was aborted part way through.

**One of the two following optional arguments can be specified.**

`phenotype_inclusion_file` (optional): Full file path to a txt file containing single column containing full PheWAS_ID of phenotypes that will be included. Cannot be used with phenotype_exclusion_file argument.

`phenotype_exclusion_file` (optional): Full file path to a txt file containing single column containing full PheWAS_ID of phenotypes that will be excluded. Cannot be used with phenotype_inclusion_file argument.

**Input**
This script accepts the following files as input:

1. covariate: A tab-separated file that contains row one of data per individual to be analysed and one column for each covariate to be included in the association model. Must start with the columns '#FID' and 'IID' both of which should be participants IDs.
2. phenotype_files: One of any number of files that contains one row of data per individual in the relevant sub-group and one column for each phenotype. For example, one such file could contain the phenotypes defined on individuals inferred as being of European descent.
3. variants_for_association: A genetic data file (in .pgen / PLINK 2.0 binary format) for the SNPs of interest.
4. alternative_manifest.csv: An alternative PheWAS manifest to that provided with the Platform.
5. phenotype_inclusion_file: A plain text file containing a single column labelled 'PheWAS_ID'. Only phenotypes with the corresponding PheWAS_IDs will be included in the analysis.
6. phenotype_exclusion_file: A plain text file containing a single column labelled 'PheWAS_ID'. Phenotypes with the corresponding PheWAS_IDs will be excluded from the analysis.

**Output**
An R object of lists. Each list is a data frame of the association results directly from Plink for each grouping variable, classically ancestry. Use `05_tables_graphs.R` to extract tables and graphs from the results.

**Examples**
1. With only required arguments specified:

```
03a_PLINK_association_testing.R /

--analysis_folder /path/to/output_folder/ /

--covariate /path/to/covariates/file.txt /

--phenotype_files /path/to/phenotypes/EUR.gz,/path/to/phenotypes/AFR.gz,
/path/to/phenotypes/SAS.gz,/path/to/phenotypes/EAS.gz /

--variants_for_association /path/to/variants_for_association /
```

```
--analysis_name my_analysis
```

2. With all arguments specified:

```
03a_PLINK_association_testing.R /
--analysis_folder /path/to/output_folder/ /
--covariate /path/to/covariates/file.txt /
--phenotype_files /path/to/phenotypes/EUR.gz,/path/to/phenotypes/AFR.gz,
/path/to/phenotypes/SAS.gz,/path/to/phenotypes/EAS.gz /
--variants_for_association /path/to/variants_for_association /
--analysis_name my_analysis \
--group_name_overide European,African,SouthAsian,EastAsian /
--PheWAS_manifest_overide /path/to/alternative_manifest/file.txt /
--plink_exe /path/to/plink2_executable /
--save_plink_tables /
--split_group European /
--N_quant_split 150 /
--N_binary_split 40 /
--model recessive /
--phenotype_inclusion_file /path/to/phenotype_inclusion_file
```

```
3. Running using split_analysis assumes already have split at least one
group, remaining arguments using default values.
```

```
03a_PLINK_association_testing.R /
--analysis_folder /path/to/output_folder/ /
--covariate /path/to/covariates/file.txt /
--phenotype_files /path/to/split_phenotypes /
--analysis_name my_analysis \
--split_analysis
```

#### 03b_R_association_testing.R

**Overview**

This script is an alternative to the `03a_PLINK_association_testing.R` script for running association analyses using R. While not streamlined for large-scale PheWAS, this script offers greater flexibility in the kind of analyses that can be run, such as fitting non-standard models or if the genetic data are not in a format readable by standard software. Further, this script allows for analyses of genomic risk scores (GRS), polygenic risk score (PRS), and multiallelic copy number variants (CNV).

GRS/PRS is inputted via a csv file with trait,group,phenotype_data,genetic_data,variable this acts as a map for the functions. See user guide for full description of this input but in brief, it allows the user to input any number of traits that may be analysed in any of the availble groupings (such as ancestry) and point to the location of the appropriate genetic and phenotype files. GRS/PRS data is analysed per-trait-group combination, results are saved as a R list object per trait that contain all trait-group combinations, this allows multiple traits to be analysed across multiple groups in a single input.

Non-GRS/PRS data is assumed to be an alternative genetic measurement with a column of IDs followed by 1-n columns of genetic data, example CNVs. Results are per column of the input file (minus ID column) with a single R object saved containing a list of results per-group such that each non-ID column will be tested for association with all available phenotypes in each of the inputted groups. Unlike the GRS/PRS input the full location of all the phenotype files is required as input.

If a covariate file is included then it must contain a column age, when selecting covariates note currently only participants with full covariate data will be analysed. Other options perform filtering on the phenotypes that are analysed, by default all phenotypes that are held in the respective phenotype tables created by `01_phenotype_preparation.R` are analysed. Minimum case numbers are options to allow the input of genetic variables that do not fully overlap with the samples used for making the phenotypes.

**Usage**

1. If testing association with a genetic risk score(s):

```
03b_R_association_testing.R (--analysis_folder=<folder> --
GRS_input=<FILE>) [--covariates=<FILE> --N_cores=<number> --
PheWAS_manifest_overide=<FILE> --binary_Case_N=<number> --
quantitative_Case_N=<number>] --phenotype_inclusion_file=<FILE> | --
phenotype_exclusion_file=<FILE>]
```

2. If testing some other association (such as with multiallelic CNVs):

```
03b_R_association_testing.R --analysis_folder=<folder> --
non_GRS_data=<FILE> --phenotype_files=<FILE> --analysis_name=<name> [--
covariates=<FILE> --group_name_overide=<text> --N_cores=<number> --
PheWAS_manifest_overide=<FILE> --binary_Case_N=<number> --
quantitative_Case_N=<number>] [--phenotype_inclusion_file=<FILE> | --
phenotype_exclusion_file=<FILE>]
```

**Arguments**

`analysis_folder` (required): Full path of the directory in which the association results will be saved.

**One of the following two arguments should be specified.**

`GRS_input` (required): Full file path of the GRS_input csv file.

`non_GRS_data` (required): Full path of the non-GRS genetic data.

`phenotype_files` (required): Comma-separated list containing the full paths of the phenotype data.

`analysis_name` (required): Name for the analysis, is used later in saving tables, so should distignuish between other analyses. For GRS analysis this name is always the trait being analysed.

`covariates` (optional): Full path of a file containing covariate data to be used in model adjustment.

`group_name_overide` (optional): Comma-separated list containing alternative group names. By default, group names are extracted from the suffix of the file names provided in the `phenotype_files` argument. For example, if a file named /home/phenotypes/EUR_phenotypes.csv were provided, the corresponding group name would be "EUR". This argument allows for a different group name to be specified. The order of names provided should match the files specified in the `phenotype_files` argument.

`N_cores` (optional): Number of cores requested if parallel computing is desired. Defaults to single core computing.

`PheWAS_manifest_overide` (optional): Full file path of the alternative PheWAS_manifest file.

`binary_Case_N` (optional): Number that represents the minimum number of cases for binary phenotype inclusion, defaults to 50.

`quantitative_Case_N` (optional): Number that represents the minimum number of cases for quantitative phenotype inclusion, defaults to 100.

**One of the two following optional arguments can be specified.**

`phenotype_inclusion_file` (optional): Full file path to a txt file containing single column containing full PheWAS_ID of phenotypes that will be included. Cannot be used with phenotype_exclusion_file argument.

`phenotype_exclusion_file` (optional): Full file path to a txt file containing single column containing full PheWAS_ID of phenotypes that will be excluded. Cannot be used with phenotype_inclusion_file argument.

**Input**

This script accepts the following files as input:

1. GRS_input: A plain text file contains the following 6 columns. A template can be found within extdata/GRS_PRS_template.csv.gz within the downloaded package.
   **trait**: Name of the trait that the GRS/PRS has been created from.
   **group:** Grouping variable that the analysis is subset to.
   **phenotype_data**: Full file path for the phenotype data that will be used in the association analysis.
   **genetic_data**: Full file path to the genetic data for the GRS/PRS.
   **column_name:** Column name in the genetic data file containing the relevant GRS genetic data.
2. non_GRS_data: A plain text file file that contains one column of participant identifiers and any number of additional columns that describe "non-GRS" phenotypes.
3. phenotype_files: One of any number of files that contains one row of data per individual in the relevant sub-group and one column for each phenotype. For example, one such file could contain the phenotypes defined on individuals inferred as being of European descent.
4. covariates: A tab-separated file that contains row one of data per individual to be analysed and one column for each covariate to be included in the association model. ID column should be '**eid**'
5. alternative_manifest.csv: An alternative PheWAS manifest to that provided with the Platform.

6. phenotype_inclusion_file: A plain text file containing a single column labelled 'PheWAS_ID'. Only phenotypes with the corresponding PheWAS_IDs will be included in the analysis.
7. phenotype_exclusion_file: A plain text file containing a single column labelled 'PheWAS_ID'. Phenotypes with the corresponding PheWAS_IDs will be excluded from the analysis.

**Output**
An R object of lists. Each list is a data frame of the association results directly from Plink for each grouping variable, classically ancestry. Use `05_tables_graphs.R` to extract tables and graphs from the results.

**Examples**
1. With only required arguments specified:

```
03b_R_association_testing.R \

--analysis_folder /path/to/output_folder/ \

--GRS_input /path/to/GRS_data/file.txt
```

**OR**

```
03b_R_association_testing.R \

--analysis_folder/path/to/output_folder/ \

--non_GRS_data /path/to/non-GRS_data/file.txt \

--phenotype_files /path/to/phenotypes/EUR.gz,/path/to/phenotypes/AFR.gz,
/path/to/phenotypes/SAS.gz,/path/to/phenotypes/EAS.gz \

--analysis_name my_analysis
```

2. With all arguments specified:

```
03b_R_association_testing.R \

--analysis_folder /path/to/output_folder/ \

--GRS_input /path/to/GRS_data/file.txt

--covariates /path/to/covariates/file.txt \

--N_cores 12 \

--phenotype_exclusion_file /path/to/phenotype_exclusion_file \

--PheWAS_manifest_overide /path/to/alternative_manifest/file.txt \

--binary_Case_N 30

--quantitative_Case_N 90
```

**OR**

```
03b_R_association_testing.R \

--analysis_folder/path/to/output_folder/ \

--non_GRS_data /path/to/non-GRS_data/file.txt \
```

```
--phenotype_files /path/to/phenotypes/EUR.gz,/path/to/phenotypes/AFR.gz,
/path/to/phenotypes/SAS.gz,/path/to/phenotypes/EAS.gz \

--analysis_name my_analysis \

--covariates /path/to/covariates/file.txt \

--group_name_overide European,African,SouthAsian,EastAsian \

--N_cores 12 \

--phenotype_overide /path/to/phenotype_subset/file.txt \

--PheWAS_manifest_overide /path/to/alternative_manifest/file.txt \

--binary_Case_N 30 \

--quantitative_Case_N 90
```

#### 04_combine_split.R

**Overview**
This script combines the results from the split_group split_analysis arguments from `03a_PLINK_assoication_testing.R` into a single result file and appends the original results file first removing the empty list and then adding the split group into it.

**Usage**:
`04_combine_split.R (--split_plink_results_folder=<folder>  --results_RDS_file=<FILE> --group_name=<name> --analysis_name=<name>)`

**Arguments**
`split_plink_results_folder` (required): Full path of the folder containing all of the PLINK results from the split_group, split_analysis option in `03a_PLINK_association_testing.R`.

`results_RDS_file` (required): Full path of the results of the analysis from the `03a_PLINK_association_testing.R` script. Is an R object.

`group_name` (required): Name of the group that was split that the results represent.

`analysis_name` (required): Name for the analysis to be used in the naming of the output files, use the same as in 03a_PLINK_association_analysis.

**Input**
This script uses the following files as input:

1. main_results_file: the output of the 03a_PLINK_association_testing.R script when not using split_analysis argument. A list of data-frames combining the results per grouping variable inputted.

**Output**
An amended main_results_file with the split group now combined into a single data-frame added to the list of data-frames.

**Example**
```
--split_plink_results_folder /path/to/split_results_folder/ \

--results_RDS_file /path/to/results_RDS_file \

--group_name EUR \
```

```
--analysis_name test_split_analysis
```

#### 05_tables_graphs.R

**Overview**

Creates result tables and graphs for the association analyses from either the GRS or plink methods. The two usages below represent inputting data from 03a_PLINK_association_testing.R and 03b_R_association_testing.R.

To produce graphs input the per_group_name_graph, per_snp_graph or R_association_graph depending on result source and transformation of the data required. There are many options to select related to filtering for MAC in plink results and general appearance of the graphs. By using the provided filter inputs it is possible to edit any individual graphs using the array of options.

**Usage**

1. If the association analysis was conducted using PLINK 2.0:

```
05_tables_graphs.R (--results_file=<FILE> --analysis_name=<name> --
plink_results --SNP_list=<FILE> --save_folder=<FOLDER>) [--
group_filter=<text> --PheWAS_ID_filter=<FILE> --
PheWAS_manifest_overide=<FILE> --max_pheno=<number> --sig_FDR=<number> --
no_save_table_all_results --no_graph_all --no_graph_sig --
max_FDR_graph=<number> --SNP_filter=<FILE> --group_name_filter=<FILE> --
save_raw_plink --MAC=<number> --MAC_case=<number> --MAC_control=<number> -
-per_group_name_graph --per_snp_graph --save_table_per_group_name --
save_table_per_snp --sex_split]
```

2. If the association analysis was conducted using R:

```
05_tables_graphs.R (--results_file=<FILE> --analysis_name=<name> --
R_association_results --save_folder=<FOLDER>) [--group_filter=<text> --
PheWAS_ID_filter=<FILE> --PheWAS_manifest_overide=<FILE> --
max_pheno=<number> --sig_FDR=<number> --no_save_table_all_results --
no_graph_all --no_graph_sig --max_FDR_graph=<number> --R_association_graph
--sex_split]
```

**Arguments**

results_file (<u>required</u>): Full file path of the results file RDS R list object.

save_folder (required): Full file path of the folder to which the output will be saved.

**If the association analysis was conducted using PLINK 2.0, the following two arguments must be specified.**

plink_results (<u>required</u>): Select if results are from 03a_PLINK_association_testing.R

SNP_list (<u>required</u>): Full path of the SNP_list file used in 02_extracting_snps.R.

**If the association analysis was conducted using R, the following argument must be specified.**

R_association_results (<u>required</u>): Select if results are from 03b_R_association_testing.R.

*Options*

group_filter (optional): Comma-separated text input, used to filter the group to which the table and graph functions are applied. Inputted groups are the ones that are retained for analysis, group here refers to the grouping variable used to subset the analysis classically ancestry.

PheWAS_ID_filter (optional): Full path of a file describing the subset of PheWAS IDs that will be included in the output.

PheWAS_manifest_overide (optional): Full file path of the alternative PheWAS_manifest file.

max_pheno (optional): Manual override for inputting maximum phenotypes analysed. Used for calculating FDR. The default used the largest number of associations in the results file of any grouping.

sig_FDR (optional): Threshold for defining a significant association based on FDR. Default is 0.01 (1%).

no_save_table_all_results (optional): Specify whether a table containing all the results should be saved (FALSE) or not (TRUE). Default is FALSE.

no_graph_all (optional): Specify whether a figure of all the results should be produced (FALSE) or not (TRUE). Default is FALSE.

no_graph_sig (optional): Specify whether a figure of the significant results should be produced (FALSE) or not (TRUE). Default is FALSE.

max_FDR_graph (optional): Maximum value for the *y*-axis to be used when the FDR for a particular association is exceptionally small. Default is 300.

R_association_graph (optional): Specify whether a figure of the results from the association analysis from 03b_R_association_testing.R should be produced (TRUE) or not (FALSE). Default is FALSE.

SNP_filter (optional): Full path of a file containing SNP IDs, used to filter the output to a given subset of SNPs. Use if wanting to apply the table and/or graphing functions a subset of results. Only works for plink_results.

group_name_filter (optional): Full path of the file containing a list of group_names that correspond with those defined in SNP_list. This can be used if the table and/or graphing functions should be applied to a subset of SNPs, such as only those in a given credible set. This option can only be used with association results generated by 03a_PLINK_association_testing.R.

save_raw_plink (optional): Specify whether the unfiltered association results from the PLINK analysis should be saved (TRUE) or not (FALSE). Default is FALSE.

MAC (optional): MAC (minor allele count) filter applied to all associations, only applicable in results from 03a_PLINK_association_testing.R Default is 20.

MAC_case (optional): Minor allele count among cases for filtering the association results. Only applicable in results from 03a_PLINK_association_testing.R and only in binary phenotypes. Default is 5.

MAC_control (optional): Minor allele count among controls for filtering the association results. Only applicable in results from 03a_PLINK_association_testing.R and only in binary phenotypes. Default is 10.

`per_group_name_graph` (optional): Select if wanting to produce graphs per-group_name. This is used when looking to report the most significant finding across several SNPs for a single construct, potentially and gene or a sentinel SNP with a credible set. It is a column in the SNP_list file. Default is FALSE.

`per_snp_graph` (optional): Specify whether a figure is produced for every SNP provided (TRUE) or not (FALSE). Default is FALSE.

`save_table_per_group_name` (optional): Select if wanting to produce tables per-group_name. This is used when looking to report the most significant finding across several SNPs for a single construct, potentially and gene or a sentinel SNP with a credible set. It is a column in the SNP_list file. Default is FALSE.

`save_table_per_snp` (optional): Specify whether a results table should be generated for every SNP provided (TRUE) or not (FALSE). Default is FALSE. Will be saved in a created folder named /analysis_name_group_per_SNP_tables. Example if the group was groupA and analysis_name top_SNPs the folder would be /top_SNPs_groupA_per_SNP_tables.

`sex_split` (optional): Specify whether the figure should be stratified by sex (TRUE) or not (FALSE). If TRUE, this will produce separate figures for males, females and combined. Default is FALSE. Does not create split tables.

**Input**
This script accepts the following files as input:

1. **results_file**: An RDS object outputted from either `03a_PLINK_association_testing.R` or `03b_R_association_testing.R.`
2. **SNP_list**: SNP_list file used in 02_`extracting_snps.R`
3. PheWAS_ID_filter: A plain text file containing a single column that lists the PheWAS IDs to be excluded from the output, must include full PheWAS_ID if wanting to exclude generated age_of_onset or sex stratified (_male and _female) versions of phenotypes created in `01_phenotype_preparation.R.`
4. alternative_manifest.csv: An alternative PheWAS manifest to that provided with the Platform.

**Output**
This script can output the following files dependent on options:

Tables

Results tables from **`03a_PLINK_association_testing.R`** data, have the following columns.

- **group**: Grouping variable used to subset the analysis, classically this will be ancestry.
- **collective_name**: Name taken from the SNP_list group_name input. Used to identify analysis and also group together multiple SNPs for per-grouping_name analysis.
- **PheWAS_ID**: Phenotype ID.
- **category**: type of phenotype, indicates how it was made and what sort of data it used.
- **description**: description of the phenotype.
- **N_ID**: Total number of participants in the analysis.
- **rsid**: SNP identifier.
- **FDR**: False discovery rate.
- **P**: P value.
- **OR**: Odds ratio.

- **Beta**: Raw Beta value from the analysis.
- **L95**: lower 95% confidence interval.
- **U95**: upeer 95% confidence interval.
- **coded_allele**: the allele which the direction of effect is based (the tested allele).
- **non_coded_allele**: the other allele.
- **minor_allele**: the allele with the lowest count/frequency of the two.
- **MAF**: Minor allele frequency.
- **MAC**: Minor allele count.
- **MAC_cases**: the minor allele count in cases only.
- **MAC_controls**: the minor allele count in controls only.
- **chromosome**: The chromosome of the SNP.
- **position**: The genomic position of the SNP.
- **Z_T_STAT**: The Z or T stat (depending on binary or quantitative phenotype).
- **SE**: Standard error.
- **effect_direction**: The direction of effect (positive or negative)
- **phenotype_group**: Major grouping of phenotype used for both graphs.
- **phenoytpe_group_narrow**: More specific grouping only used in tables.
- **short_desc**: The short description used in the graphs rather than full description seen in the table.
- **Info_score**: INFO score shows the confidence of the call of the SNP if it was imputed.
- **firth**: Whether Firth regression was used by Plink.
- **TEST**: The genetic model used.
- **Error_flag**: the recorded error flag from PLINK.
- **ID**: the created ID of the SNP used by appending the rsid and with the two alleles in alphabetical order.
- **graph_save_name**: the save name for the graph produced from the SNP taken from SNP_list.
- **sex_pheno_identifier**: identifies whether the phenotype is sex stratified.

Results tables from **`03b_R_association_testing.R`** data, have the following columns.

- **name**: Name of the analysis, for GRS/PRS refers to the trait for other inputs refers to the name given to the specific column the data is stored under used when running `03b_R_association_testing.R.`
- **PheWAS_ID**: Phenotype ID.
- **category**: type of phenotype, indicates how it was made and what sort of data it used.
- **phenotype**: description of the phenotype.
- **FDR**: False discovery rate.
- **P**: P value.
- **OR**: Odds ratio.
- **Beta**: Raw Beta value from the analysis.
- **L95**: lower 95% confidence interval.
- **U95**: upeer 95% confidence interval.
- **SE**: Standard error.
- **phenotype_group**: Major grouping of phenotype used for both graphs.
- **phenoytpe_group_narrow**: More specific grouping only used in tables.

- **short_desc**: The short description used in the graphs rather than full description seen in the table.
- **effect_direction**: The direction of effect (positive or negative)
- **group**: Grouping variable used to subset the analysis, classically this will be ancestry.
- **name_group**: concatenated name and group, used for file saving.
- **table_save**: used for non GRS data in saving files.
- **sex_pheno_identifier**: identifies whether the phenotype is sex stratified.

Graphs
Two graphs can be produced

- suffix for the save name **all_pheno** produces a graph showing all the association results. The Y axis is -log$^{10}$(FDR) with a line of significance in red. The x-axis has the **phenotype_group** and orders graph by the lowest association within each group and then in descending order per group. The direction of effect is signified by a triangle either pointing up or down. Phenotypes that have associations deemed significant are labelled using the short_desc within the tables. The value in parenthesis is the **category** of phenotype seen in the tables, this also indicates whether a phenotype is male stratified (male), female stratified (female) or age of onset (age_of_onset).
- suffix for the save name **sig_pheno** produces a graph showing only associations deemed significant. The Y axis is -log$^{10}$(FDR) with a line of significance in red. The x-axis has individual phenotypes ordered by **phenotype_group** the lowest association within each group and then in descending order per group (only of significant results). Phenotypes are labelled using **short_desc.** The value in parenthesis is the **category** of phenotype seen in the tables, this also indicates whether a phenotype is male stratified (male), female stratified (female) or age of onset (age_of_onset). The direction of effect is signified by a triangle either pointing up or down. The **phenotype_group** is shown in the legend under Phenotypic Category.

**Examples**
1. With only required arguments specified:

```
05_tables_graphs.R \
--results_file /path/to/association_results/file.gz \
--plink_results \
--save_folder /path/to/save_folder/ \
--SNP_list /path/to/SNP_list
```

**OR**

```
05_tables_graphs.R \
--results_file /path/to/association_results/file.gz \
--save_folder /path/to/save_folder/ \
--R_association_results
```

2. With some options specified:

```
05_tables_graphs.R \
--results_file /path/to/association_results/file.gz \
```

```
--save_folder /path/to/save_folder/ \
--plink_results \
--SNP_list /path/to/SNP_list \
--max_pheno 1000 \
--sig_FDR 0.05 \
--MAC 30 \
--sex_split
```

**OR**

```
05_tables_graphs.R \
--results_file /path/to/association_results/file.gz \
--save_folder /path/to/save_folder/ \
--R_association_results \
--save_folder /path/to/alternative/save_folder/ \
--max_pheno 1000 \
--sig_FDR 0.05 \
--R_association_graph \
--sex_split
```
