## Supplementary Methods and Figures for "DeepPheWAS: an R package for phenotype generation and association analysis for phenome-wide association studies"

### Supplementary material

#### Supplementary methods

DeepPheWAS is an R package for PheWAS analysis using a series of function accessed through R scripts optimised for, but not exclusively applicable to, UK Biobank data. The functions separate data wrangling, phenotype generation and association analysis into discrete steps. By separating out the analysis the platform provides capacity for customisation to meet user requirements. Here we describe how the key features of the platform work. For a detailed guide to each script in the package, files formats and the various input options, see the **User Guide**.

##### Data wrangling

The first group of functions convert the data into a format that the remaining functions can analyse; this is performed through two scripts. The first script (01_minimum_data.R) creates a file that contains all required columns of non-linked healthcare or genetic data. In UK Biobank this is questionnaire or measurements undertaken by the UK Biobank study. A list of column names is provided to guide the extraction and can be edited for column names of non-UK Biobank studies. The second script (02_data_preparation.R) creates a file that contains health records from linked datasets alongside study-acquired data on self-reported illness and operations into a single long file. This script is specific for UK Biobank, but non-UK Biobank studies can format their data to match the output of this script. Formats of both files can be found in the user guide.

##### Phenotype generation

There are six categories of phenotype that are created by DeepPheWAS. These are grouped into categories depending on the type of data they use, and the methods used to generate them. The six categories are: phecode phenotypes; Data-Field phenotypes; Combined Data-Field phenotypes; formula phenotypes, primary care quantitative phenotypes and composite phenotypes. Alongside these six types of phenotypes, DeepPheWAS also creates two kinds of concept, concepts can either be code-list-concepts or composite concepts, explained further below. Every phenotype/concept described below will contain the participant ID, number of codes/value and earliest date of code/value.

#### Phecode phenotypes

Phecodes are a coding ontology developed by mapping ICD-9 codes into 1867 clinically relevant groupings (Denny *et al.*, 2013); these maps were later extended to ICD-10 codes (Wu *et al.*, 2019). In addition to these primary groupings there are also 233 distinct groups of exclusions that encompass one or more phecodes. These groupings, we call range-IDs, are used to exclude controls based on similarity to the cases, for example participants with type 2 diabetes may be excluded from being controls in a study of type 1 diabetes given some phenotypic similarity and potential for misclassification of controls. DeepPheWAS creates the 1848 phenotypes using the phecode maps for ICD-9 and ICD-10 data using the 03a_phecode_generation.R script. ICD codes can be extracted from any source; for UK Biobank these are hospital episode statistics (ICD-9, ICD-10), mortality data (ICD-10) and cancer registry (ICD10). Using the same functions DeepPheWAS also saves the 233 distinct range-IDs, these can be called upon for developing composite phenotypes, discussed later.

#### Data-Field phenotypes

Data-Field phenotypes are phenotypes derived from a single data field and are created using the 03b_data_field_phenotypes.R script, guided by inputs into the PheWAS-manifest file (see user guide). These can be quantitative or binary phenotypes. To create binary phenotypes from categorical data with over two responses, DeepPheWAS can collapse multiple responses into a binary set (or can ignore certain responses, where specified by the user). For quantitative phenotypes where there are multiple measures DeepPheWAS allows selection of the first, last, minimum, or maximum values, this can be edited by the user. There are 467 Data-Field ID phenotypes, 68 binary and 399 quantitative, created by DeepPheWAS.

#### Combined Data-Field phenotypes

Combined Data-Field phenotypes are quantitative Data-Field phenotypes that made by combining two or more individual Data-Field phenotypes. As for quantitative Data-Field phenotypes, the first, last, minimum, or maximum value can be extracted from the component phenotypes. Combined Data-Field phenotypes are created using the 03b_data_field_phenotypes.R script, guided by inputs into the PheWAS-manifest file. This script is employed to reduce the number of highly correlated phenotypes such as when using lateralised measures, e.g., ocular pressure in the left and right eye can be reduced to a single measure of ocular pressure. There are six Combined Data-Field phenotypes created by DeepPheWAS.

#### Primary care quantitative phenotypes

Primary care data contains standard clinical codes that indicate diagnosis, administration codes, prescription codes and measurements values for example from blood tests or physical measures such as blood pressure. Primary care quantitative phenotypes are made by extracting this measurement data. The extraction is guided by the PheWAS_manifest and lists of codes to extract stored with the package /extdata/PQP_codes/ with optional filters for minimal and maximum values. There are three primary care quantitative phenotypes created by DeepPheWAS

#### Formula phenotypes

Formula phenotypes are generated by combining data fields, clinical data or other phenotypes in ways which cannot be done using the inherent functions of DeepPheWAS. All other phenotypes can be edited outside of the R code through editing specific mapping files. However, as the formula that create formula phenotypes are hard coded into the functions they cannot currently be changed without altering the underlying code. An example of a formula phenotype is estimated glomerular filtration rate which is calculated by applying a formula to measured cystatin C combined with sex and age. There are seven formula phenotypes created by DeepPheWAS.

#### Composite phenotypes, code list concepts and composite concepts

Concepts can be code-list concepts or composite concepts. A code-list concept is a list of codes that define a single construct. This construct can be the presence or absence of a disease, a symptom, or a list of related medications. Code-list concepts can contain clinical codes which are any codes that are stored in the health_records.txt.gz generated by 02_data_preparation.R script, or prescription codes but not both. Natively for UK Biobank data, the clinical codes that are used are ICD-9, ICD-10, Read V2, Read V3, self-report non-cancer diagnosis, self-report operations and OPSC-4 (codes for operations stored within hospital records). Composite concepts are created using the same methods of composite phenotypes described below and are distinct to composite phenotypes by the lack of controls. The lists of codes included for each concept is editable, and new concepts can be created as required. Currently there are 80 code-list concepts and five composite concepts included in DeepPheWAS.

Composite phenotypes are created using tools developed for DeepPheWAS that combine any of the existing phenotypes/concepts described above by interpreting formulae, including parentheses, using Boolean operators AND, OR, NOT. The formulae are represented in a composite phenotype mapping file and can use numbers of codes/value of code alongside date relationships between codes to develop novel phenotypes. This file can be edited to generate a custom set of phenotypes for association testing. All lists of codes for clinical concepts and formulae for composite phenotypes have been developed in collaboration with at least one clinically trained individual. There are 69 composite phenotypes currently in DeepPheWAS.

The total number of phenotypes/concepts currently created by DeepPheWAS is 2486.

### Phenotype preparation

Phenotypes are prepared for association using the 01_data_preparation.R script. This first splits the population into user-defined groups (commonly ancestry) and optionally will create sex-stratified and age of onset versions of designated phenotypes. The phenotypes can then be filtered to those with a minimum number of cases (binary phenotypes) or a minimum total number of individuals (quantitative traits), as specified by the user. There is a user option to filter by relatedness followed by a further filter by case number (as removing related individuals could reduce cases sufficiently for the phenotype to be excluded), this saves the filtered phenotypes as an R object. The second step converts the saved R object created from the previous stage into tables and applies any transformation specified (currently only inverse normal transformations are accepted). At any stage phenotypes can be excluded based on user-inputted lists.

### Association testing

Two methods for association testing are provided: (i) one for any genetic variable that can be stored in a PLINK or BGEN binary file 03a_PLINK_association_testing.R; (ii) one for any instrument that cannot be analysed via PLINK2 03b_R_association_testing.Rscript.

#### PLINK analysis

##### *Genetic data extraction*

DeepPheWAS provides tools for any genetic variants that can be stored in either PLINK binary files or .bgen files, most commonly single nucleotide polymorphisms (SNPs), using the 02_extracting_SNPs.R script. Variants are extracted using PLINK2 (Chang *et al.*, 2015)or bgenix (Band and Marchini, 2018), integrated into R scripts. At minimum the user will require the installation of PLINK 2.0. The user provides a file describing the SNPs of interest, their corresponding chromosome IDs and desired coded allele (see user guide for formatting requirements). The SNPs are extracted and combined into a single .pgen format file for analysis in PLINK 2.0.

##### *Association testing*

Association testing for variants extracted with 02_extracting_SNPs.R is achieved via 03a_PLINK_association_testing.R which utilises PLINK 2.0. PLINK 2.0 was selected as it is a freely available, highly efficient method of analysing suitable genetic data, which has an in-built "Firth fall-back" option, which can prevent type 1 error in association testing with low allele counts in the presence of case-control imbalance. The user may specify one of additive, dominant, recessive and genotypic genetic models that is accepted by PLINK 2.0 and may exclude phenotypes prior to association testing. The user may also select to split the analysis to allow more efficient analysis using parallel computing. However, parallel computing is unlikely to be needed unless analysing a large number of variants in studies with a large sample size.

#### R analysis

For those genetic variables that cannot be analysed using PLINK 2.0 DeepPheWAS provides an R interface using generalised linear models (GLMs) in 03b_R_association_testing.R. As for PLINK analysis the user can select phenotypes to exclude.

### Table and graph creation

Association results are processed into tables, using the 05_tables_graphs.R script, that in the case of PLINK analysis orientate the direction of effect estimate to the inputted coded allele. For PLINK analysis, minor allele count (MAC) filters are used to prune associations. All values for the MAC filters described can be edited by the user. An overall MAC combining MAC in cases and controls larger than 20 is required.  By calculating the expected MAC in cases, we are able to apply different filter conditions on cases depending on how the observed MAC differs from the expected MAC, which may retain more association results. A MAC filter for controls is set at 10. In cases a MAC filter of 5 is applied except in certain conditions when relating the observed MAC to the expected MAC. In associations where the observed case MAC is less than 5 but greater than or equal to 1 and the expected MAC was greater than or equal to 7, we retain the association. In associations where the observed MAC is between 3 and 5 and the expected MAC is less than or equal to 1 we again retain the association. For R association results there is no such filtering process.

The false discovery rate (FDR) is calculated based on one of the highest number of associations in any of the grouped analyses (normally grouped per-ancestry) or a user-set figure (which cannot be lower than the total number of associations). Tables and Manhattan plots are produced with the filtered results. Manhattan plots for all associations can be plotted, as can plots with only significant associated phenotypes, each with various options (see user guide). Tables and graphs will be produced for all groupings (such as ancestry). False discovery rate (FDR) is used to highlight significant associations, with the default threshold set at FDR<=0.01.

### Supplementary Figures and Tables

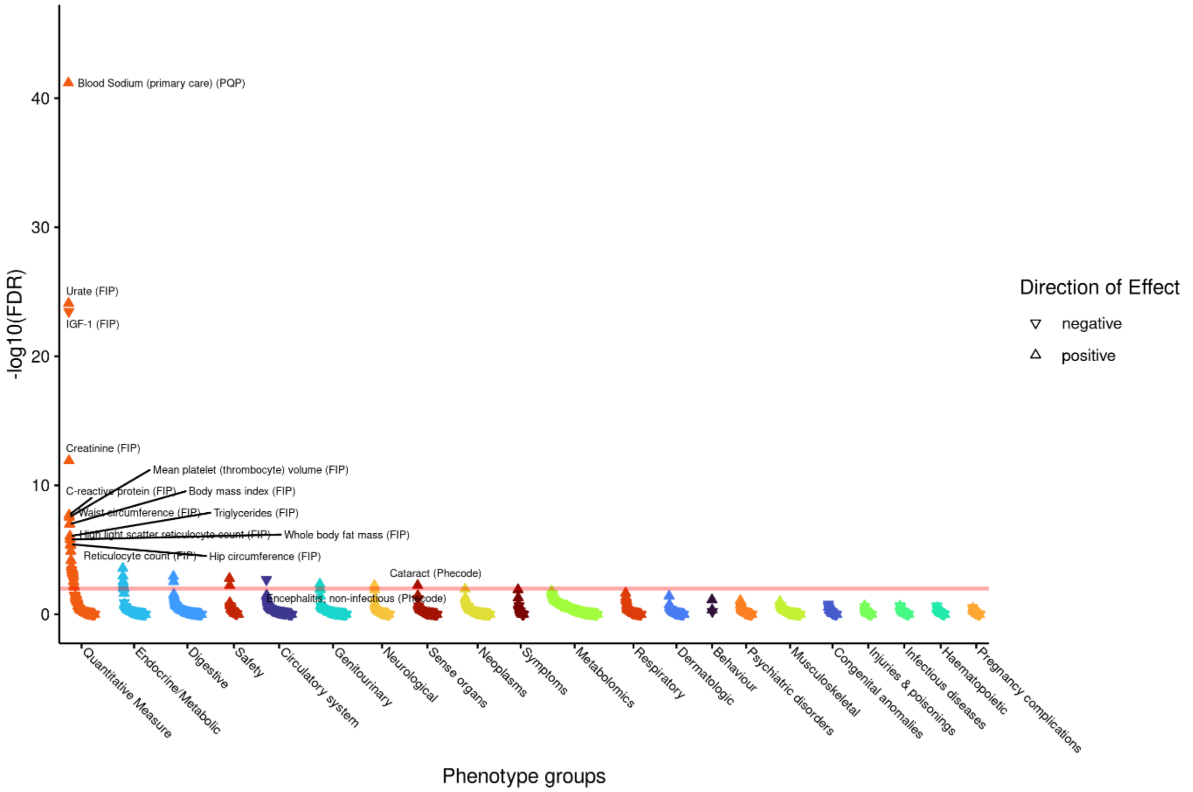

Supplementary Figure 1 - Manhattan plot of associations with rs7193778 (nearest genes *NFAT5* and *TERF2*, results orientated to C allele) under an additive genetic model grouped by phenotype group, ordered by largest association within a given group. (FDR=false discovery rate, CP=composite phenotype, FIP=Data-Field ID phenotype, FP=formula phenotype, results with an FDR of 0 are set as -log$_{10}$(FDR) of 300)

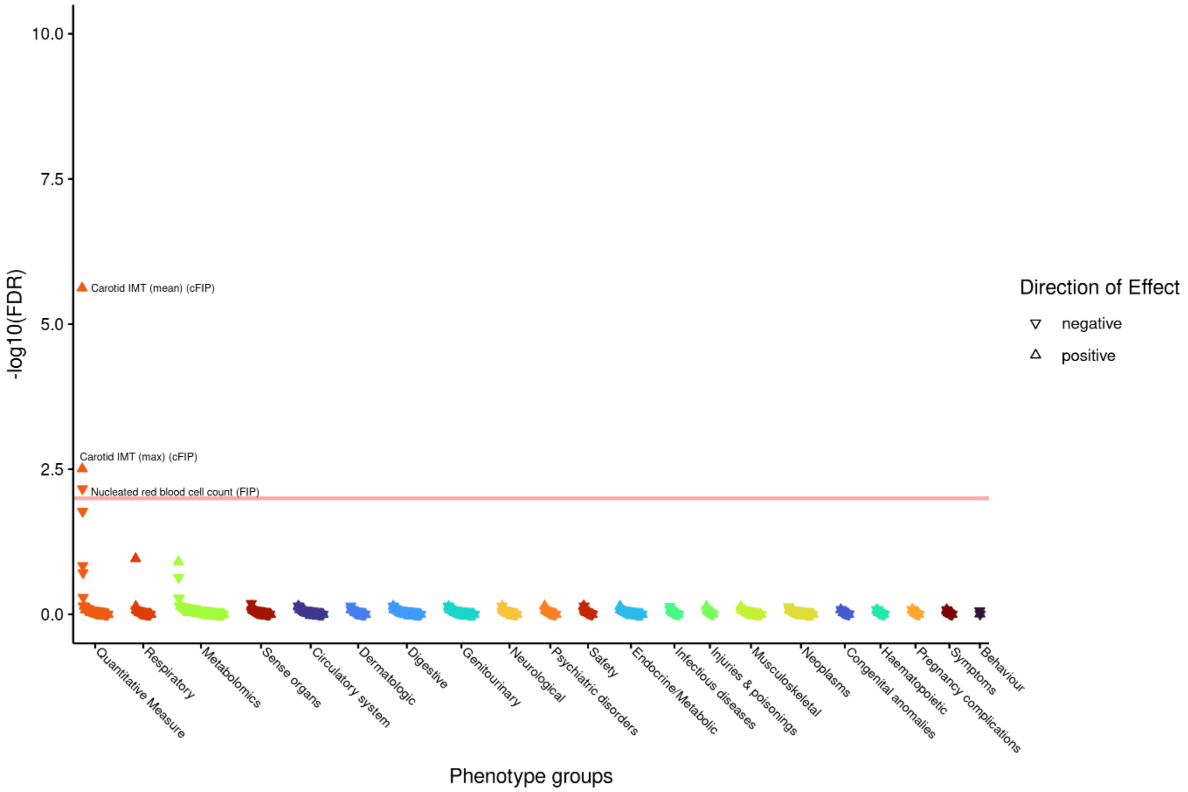

Supplementary Figure 2 - Manhattan plot of associations with rs2912062 (nearest genes *ANGPT2* and *AGPAT5,* results orientated to C allele) under an additive genetic model grouped by phenotype group, ordered by largest association within a given group. (FDR=false discovery rate, CP=composite phenotype, FIP=Data-Field ID phenotype, cFID=Combined Data-Field ID phenotype, FP=formula phenotype, results with an FDR of 0 are set as $-\log_{10}$(FDR) of 300)

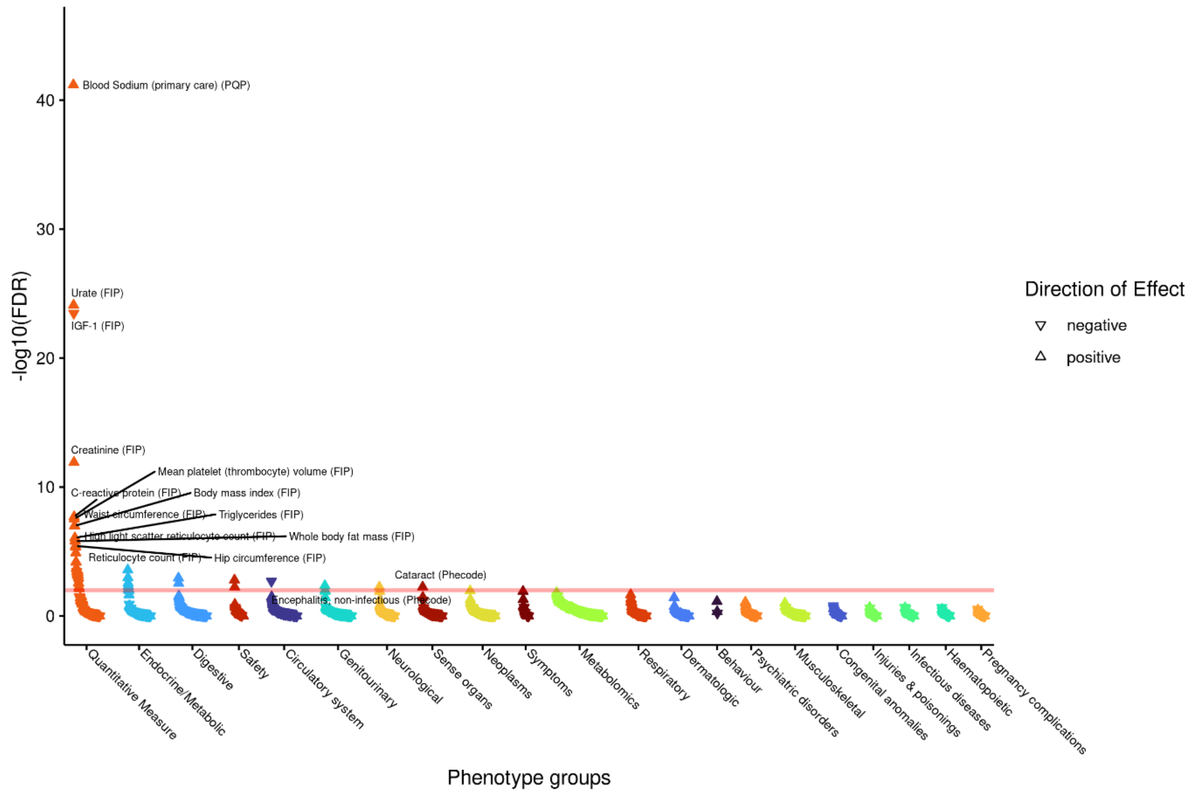

Supplementary Figure 3 - Manhattan plot of associations with SERPINA1 (rs28929474, results orientated to T allele) under an additive genetic model grouped by phenotype group, ordered by largest association within a given group. (FDR=false discovery rate, CP=composite phenotype, FIP=Data-Field-ID phenotype, FP=formula phenotype, results with an FDR of 0 are set as $-\log_{10}$(FDR) of 300)

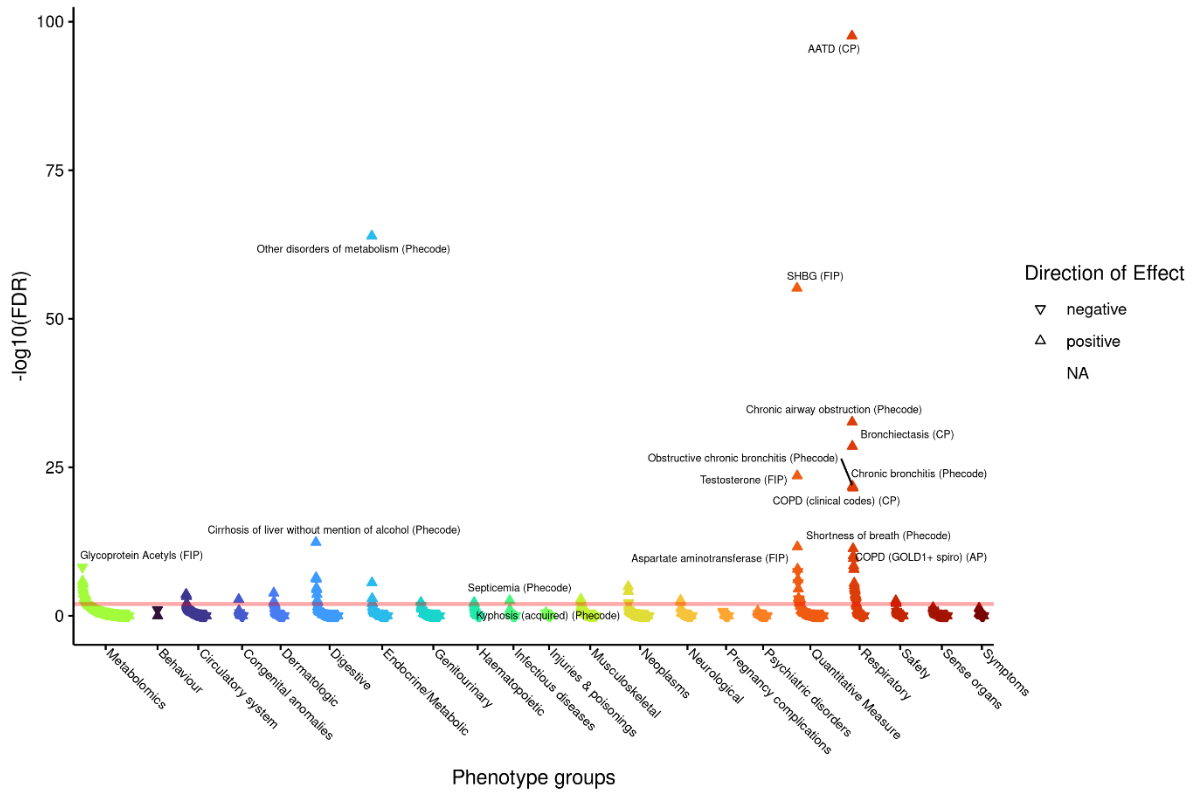

Supplementary Figure 4 - Manhattan plot of associations with *SERPINA1* (rs28929474, results orientated to T allele) under the recessive genetic model grouped by phenotype group, ordered by largest association within a given group. (FDR=false discovery rate, CP=composite phenotype, FIP=Data-Field ID phenotype, FP=formula phenotype, results with an FDR of 0 are set as -$\log_{10}$(FDR) of 300)

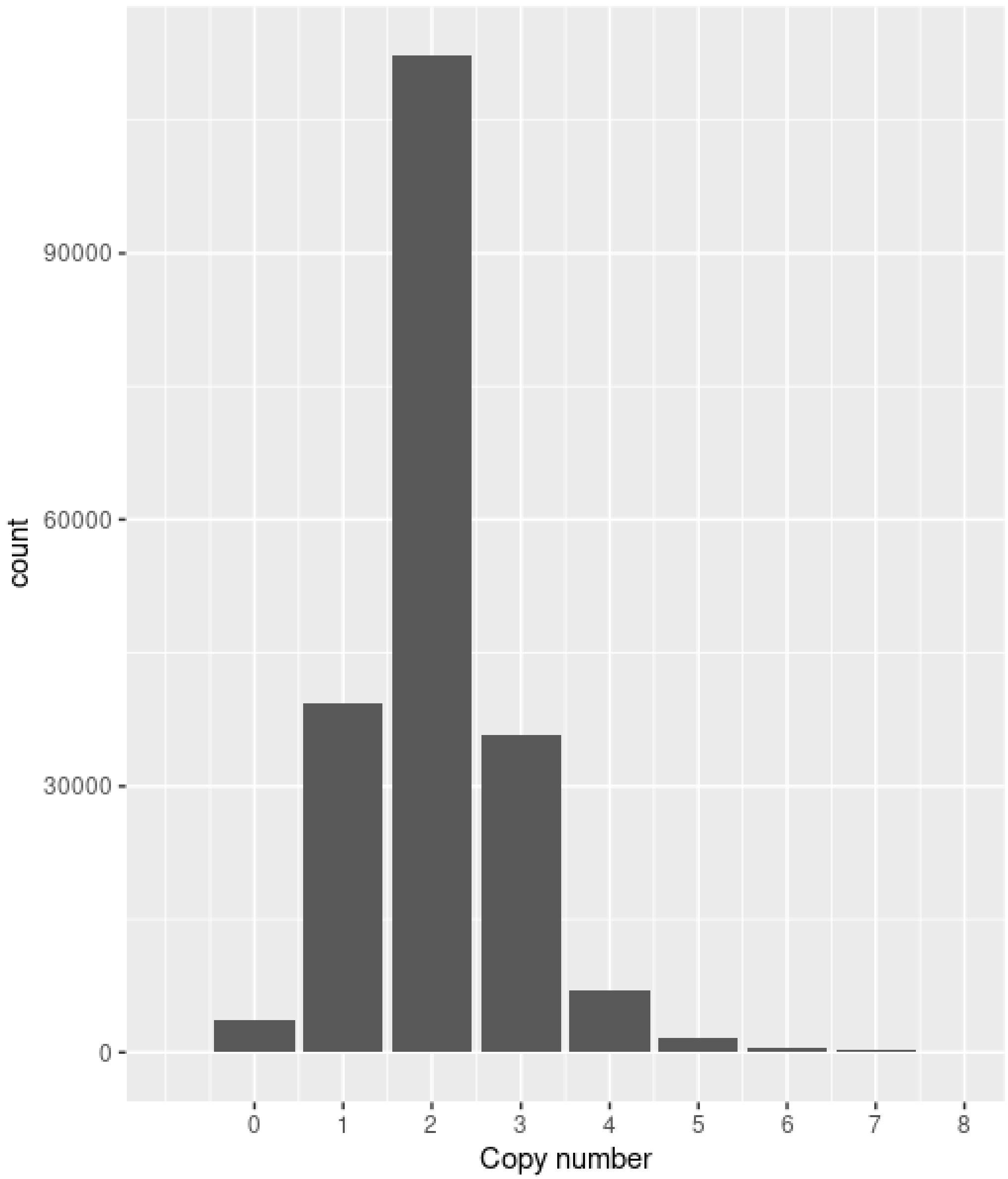

Supplementary Figure 5 – bar chart showing count of copy number in UK Biobank (N=200453) for CCL3L1

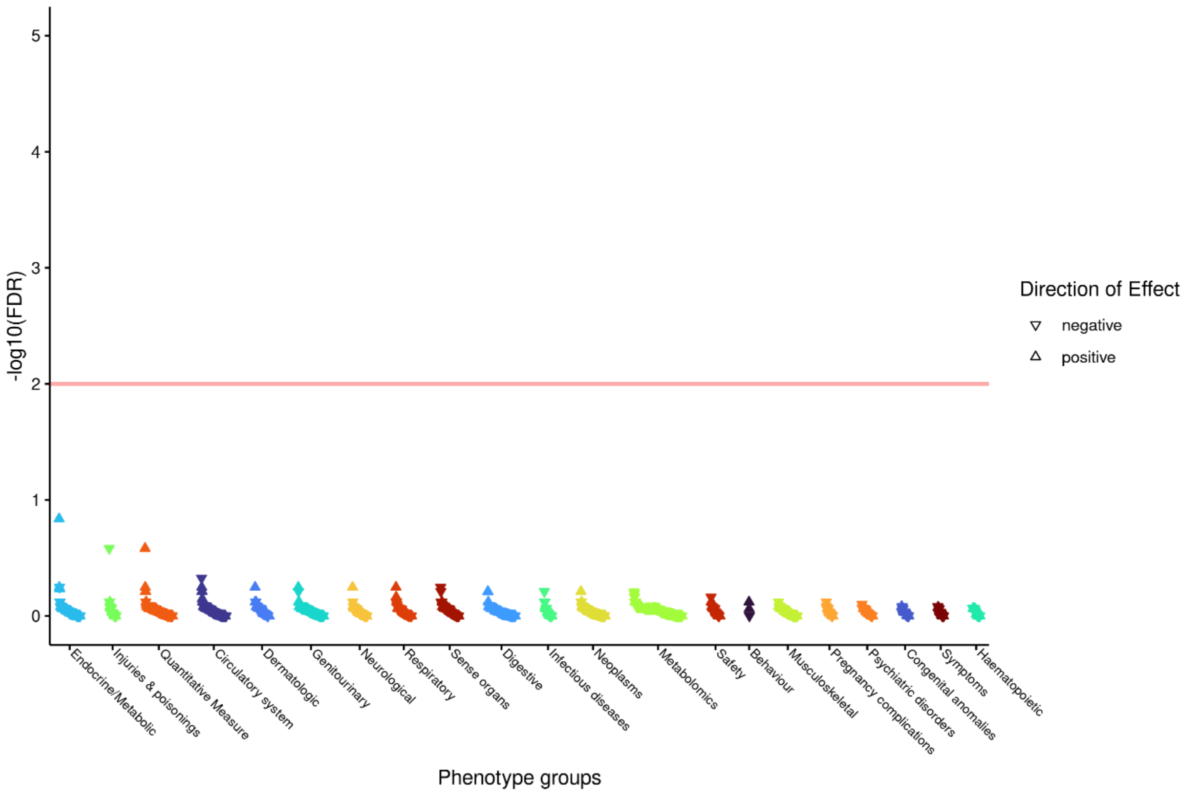

Supplementary Figure 6 – Manhattan plot of associations with *CCL3L1* copy number, grouped by phenotype group, ordered by largest association within a given group. (FDR=false discovery rate)

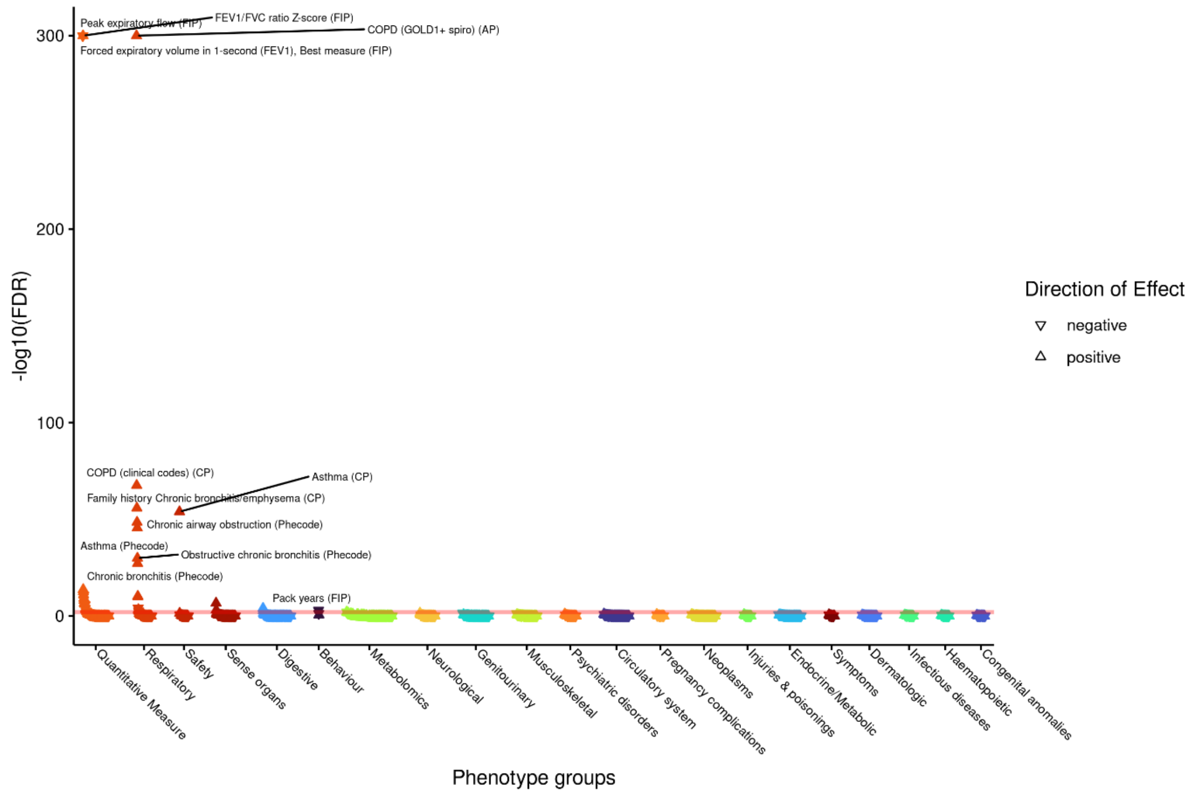

Supplementary Figure 7 – Manhattan plot of associations with GRS of lung function weighted to the ratio ($FEV_1$/FVC), grouped by phenotype group, ordered by largest association within a given group. (FDR=false discovery rate, CP=composite phenotype, FIP=Data-Field ID phenotype, FP=formula phenotype, results with an FDR of 0 are set as -$\log_{10}$(FDR) of 300)

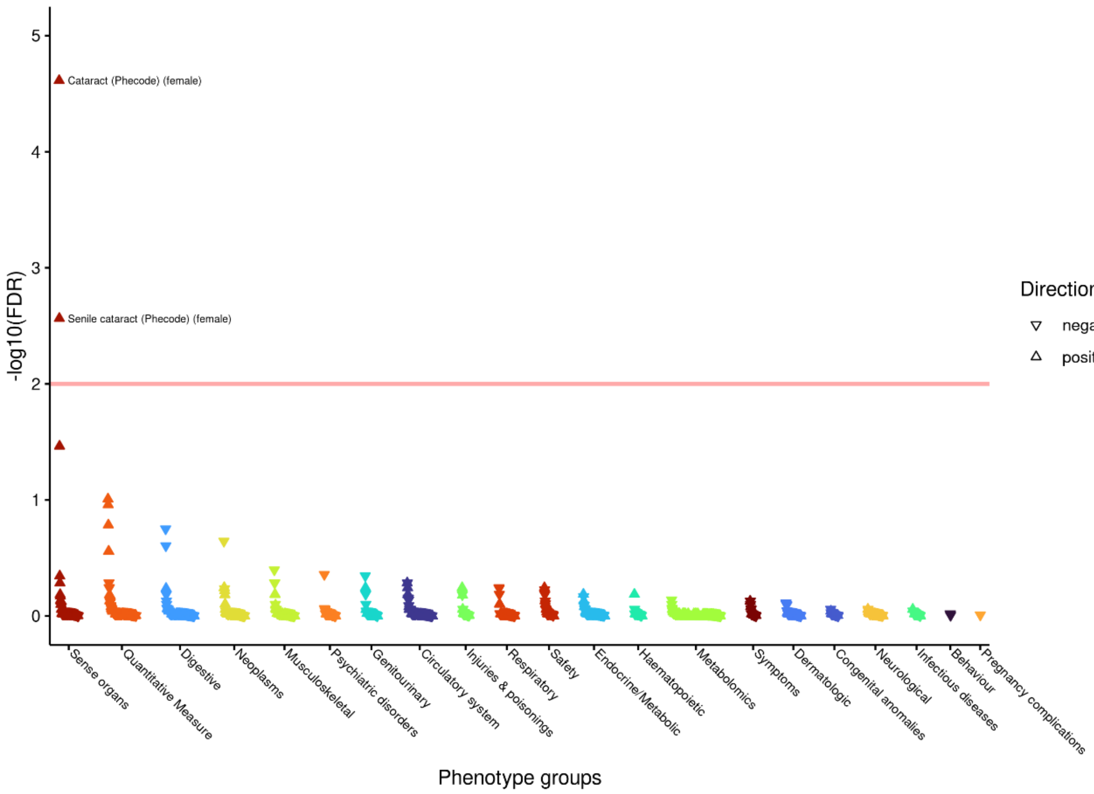

Supplementary Figure 8 - Manhattan plot of associations with rs12777332 (nearest genes *CASP7* and *NRAP*, results orientated to G allele) in females only under an additive model, grouped by phenotype group, ordered by largest association within a given group. (FDR=false discovery rate, CP=composite phenotype, FIP=*Data-Field ID* phenotype, FP=formula phenotype, results with an FDR of 0 are set as $-\log_{10}$(FDR) of 300)

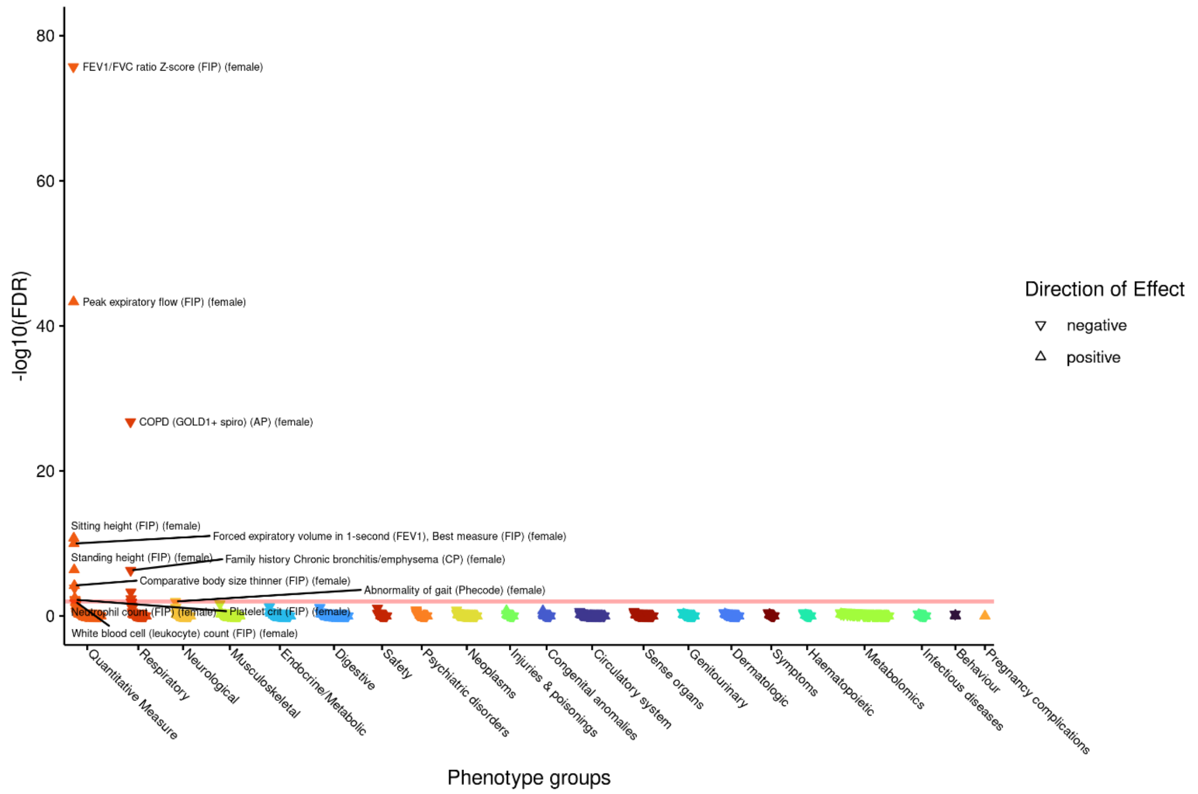

Supplementary Figure 9 - Manhattan plot of associations with rs7697189 (nearest gene *HHIP,* results orientated to C allele) in females only under an additive model, grouped by phenotype group, ordered by largest association within a given group. (FDR=false discovery rate, CP=composite phenotype, FIP=*Data-Field ID* phenotype, FP=formula phenotype, results with an FDR of 0 are set as $-\log_{10}$(FDR) of 300)

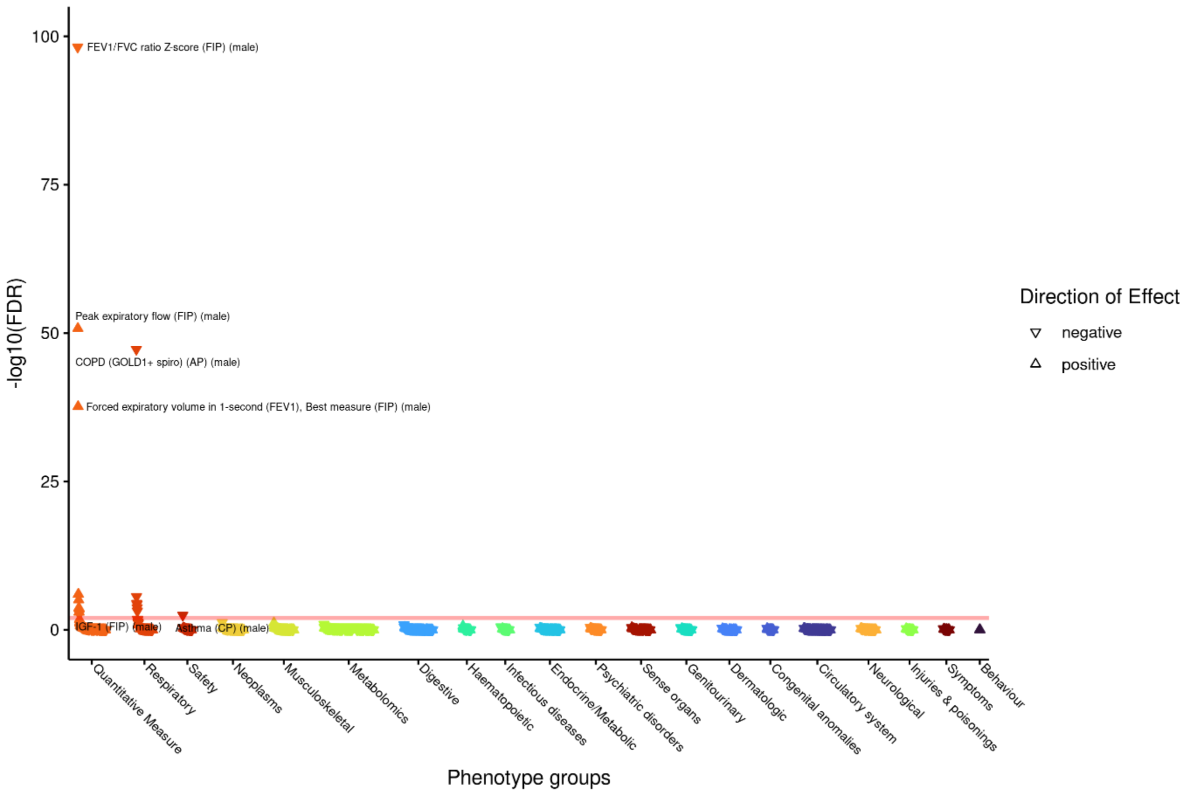

Supplementary Figure 10- Manhattan plot of associations with rs7697189 (nearest gene *HHIP,* results orientated to C allele) in males only under an additive model, grouped by phenotype group, ordered by largest association within a given group. (FDR=false discovery rate, CP=composite phenotype, FIP=*Data-Field ID* phenotype, FP=formula phenotype, results with an FDR of 0 are set as -$\log_{10}$(FDR) of 300)

Supplementary Table 1 (Excel sheet rs7193778-sodium) – results from PheWAS of rs793778 (nearest genes *NFAT5* and *TERF2,* results orientated to C allele) under an additive genetic model. (N_ID=total number of participants in analysis, FDR=false discovery rate, P=p-value, OR=odds ratio, L95=lower 95% confidence interval, U95=upper 95% confidence interval, MAF=minor allele frequency, MAC=minor allele count, Z_T_STAT=either Z/T STAT depending on binary or continuous phenotype, SE=standard error, short_description=truncation description used in figures)

Supplementary Table 2 (Excel sheet rs2912062-carotid) - results from PheWAS of rs2912062 (nearest genes *ANGPT2* and *AGPAT5,* results orientated to C allele) under an additive genetic model. (N_ID=total number of participants in analysis, FDR=false discovery rate, P=p-value, OR=odds ratio, L95=lower 95% confidence interval, U95=upper 95% confidence interval, MAF=minor allele frequency, MAC=minor allele count, Z_T_STAT=either Z/T STAT depending on binary or continuous phenotype, SE=standard error, short_description=truncation description used in figures)

Supplementary Table 3 (Excel sheet *SERPINA1*-additive) – results from PheWAS of *SERPINA1* (rs28929474, results orientated to T allele) under an additive genetic model. (N_ID=total number of participants in analysis, FDR=false discovery rate, P=p-value, OR=odds ratio, L95=lower 95% confidence interval, U95=upper 95% confidence interval, MAF=minor allele frequency, MAC=minor allele count, Z_T_STAT=either Z/T STAT depending on binary or continuous phenotype, SE=standard error, short_description=truncation description used in figures)

Supplementary Table 4 (Excel sheep *SERPINA1*-recessive) – results from PheWAS of *SERPINA1* (rs28929474, results orientated to T allele) under the recessive genetic model. (N_ID=total number of participants in analysis, FDR=false discovery rate, P=p-value, OR=odds ratio, L95=lower 95% confidence interval, U95=upper 95% confidence interval, MAF=minor allele frequency, MAC=minor allele count, Z_T_STAT=either Z/T STAT depending on binary or continuous phenotype, SE=standard error, short_description=truncation description used in figures)

Supplementary Table 5 (Excel sheet CCL3L1) – Results from PheWAS of copy number of CCL3L1. (FDR=false discovery rate, P=p-value, OR=odds ratio, L95=lower 95% confidence interval, U95=upper 95% confidence interval, SE=standard error, short_description=truncation description used in figures)

Supplementary Table 6 (Excel sheet GRS) – Results from PheWAS of lung function GRS with ratio weights. (FDR=false discovery rate, P=p-value, OR=odds ratio, L95=lower 95% confidence interval, U95=upper 95% confidence interval, SE=standard error, short_description=truncation description used in figures)

Supplementary Table 7 (Excel sheet rs12777332-CASP7) – Results from PheWAS of rs12777332 (nearest genes *CASP7* and *NRAP*, results orientated to G allele) under an additive genetic model. (FDR=false discovery rate, P=p-value, OR=odds ratio, L95=lower 95% confidence interval, U95=upper 95% confidence interval, SE=standard error, short_description=truncation description used in figures)

Supplementary Table 8 (Excel sheet rs7697189-HHIP) – Results from PheWAS of rs7697189 (nearest gene *HHIP*, results orientated to C allele) under an additive genetic model. (FDR=false discovery rate, P=p-value, OR=odds ratio, L95=lower 95% confidence interval, U95=upper 95% confidence interval, SE=standard error, short_description=truncation description used in figures)
